# Defining the pre-admission factors that modify risk of acute complications, survival and long-term recovery from COVID-19

**DOI:** 10.1101/2025.06.29.25330493

**Authors:** HE Baxendale, C Matthews, COVID19 Eastern NIHR Clinical Research Network (CRN), R Vancheeswaran, A Vuylsteke, C Denny, E Hopley, M Toshner, I Boubriak, J Newman, N Hannan, A Wilkinson, A Barlow, N Harriott, N Jones, S Webb, HICC: Humoral Immune Correlates for COVID19 (Collaborative Group/Consortium), W Schwaeble, J Heeney, M Law, D-L Couturier

## Abstract

To understand the immune correlates of protection to COVID-19, we ran a longitudinal prospective multicentre study recruiting 425 patients with COVID-19 from the start of the UK pandemic in April 2020 through the first 3 waves with study completing in 2022. Here we identify the demographic factors and pre-admission symptoms impacting on COVID-19 disease severity, acute clinical course and long-term recovery to be considered in subsequent serological analysis.

Preadmission symptom cluster and duration of symptoms associated with risk of thrombosis/renal disease and pneumothorax respectively. Renal complications were more likely to be seen in the first wave compared to the second wave of the pandemic. Survival to discharge was independently associated with ethnicity, age (<40YRSVS >60yrs) and absence of comorbidities however hazard of death was increased for all hospitalised patients who had received at least one COVID vaccine (as high-risk patient) pre-admission, had shorter time from symptom onset to admission, or had comorbidities. Most patients reported long term sequelae with neuromuscular and cognitive effects dominating early on and mood disturbance and breathlessness persisted to 12 months. Whilst COVID-19 severity score was the dominant risk factor for persistent dyspnoea, gender was the dominant risk factor for persistent mood disorder and associated with a greater risk of persistent gastrointestinal and cognitive problems.

## Introduction

Since the start of the Coronavirus-19 disease (COVID-19) pandemic, huge progress had been made in understanding the risk factors for developing and dying from severe disease. Early in the pandemic, dominant themes emerged reporting demographic factors such as gender, ethnicity, body mass, age and disease comorbidities associated with severe disease and reduced survival from acute infection {1}. Admission severity scoring systems have been developed, some of which take certain demographic features into account to inform early triage and treatment pathways. Age (>50 years), male gender and presence of comorbidities are demographic features included in some scoring {2} however there is clearly regional variation in their applicability noting the global regional variation seen in risk factors {3}, survival {4} viral variant dominance {5} and vaccination status {6} as carefully monitored by the World Health Organisation (WHO) and Our World in Data reports {7}. Applying a risk scoring system generated for one population may not be pertinent to another with a recent report of overestimation of death when a mortality scoring system developed in the UK from UK cohorts {8} was applied in Australia {9}. Local knowledge relevant to the population being treated is needed to ensure applicability.

As people recovered from acute infection, persistent and new sequelae manifest {10; 11}. These have been found to disproportionately affect certain patient groups and whilst severity of acute illness is a risk factor, several demographic factors also associated including female sex, middle age, two or more comorbidities, and more acute severe illness {12}. Global research efforts are ongoing to understand the sequelae of COVID and determine the biological mechanisms that explain them {13}. The prospective long term follow up UK study of patients hospitalised with COVID {14} (PHOSP-COVID) reports that the persistent symptoms of breathlessness are not associated with objective measures of cardiorespiratory compromise {15} and likely to be multifactorial with emotional and physical factors contributing {16}. In contrast, cognitive impairment, whilst also multifactorial, had been associated with signatures of inflammation {17).

Much of the learning about the risk factors associated with the clinical course of acute COVID is based on patients recruited early in the pandemic. There have been fewer studies evaluating the change in course with subsequent waves of infection with new viral variants and the impact of vaccination on the identified risk factors for severe acute disease and poor recovery. In addition, whilst there is now an extensive body of knowledge detailing the longer-term sequelae of COVID-19, fewer studies detailing the entire clinical course from symptom onset to long term recovery in well characterised clinical cohorts.

Here we report the clinical results forming part of the prospective longitudinal multicentre study (the Humoral Immune Correlates of Protection study (HICC) {18}, describing the demographic, clinical course and outcome to 12 months after hospital discharge in a large cohort of patients recruited across 3 waves of the UK pandemic of whom 17% received at least 1 vaccination. We identify pre-admissions risk factors that associate independent of the WHO COVID-19 Severity Classification {19} that associate with acute complications, survival and long-term recovery. These factors are then taken into account in evaluating the humoral correlates of protection to severe COVID-19 in this cohort (manuscript in preparation).

## Methods Summary

### Study Design and Approvals

This study is part of the Human Immune Correlates for COVID-19 (HICC) Study funded as an Urgent Public Health study jointly by the National Institute of Health Research (NIHR) and United Kingdom Research and Innovation (UKRI), study number: COV0170 / MC_PC_20016 {18}. It is a prospective multicentre study in England, recruiting patients hospitalised with COVID-19 over the first 3 waves of the COVID-19 pandemic and following them through the acute phase of disease and in convalescence up to 1 year. Non-hospitalised patients were also recruited representing patients with asymptomatic or mild disease. In this report, we describe the demographic of the patient cohort by disease severity, complications of acute disease survival to discharge and long-term recovery. We define the demographic risk factors for poor outcome of hospital admission and chronic sequelae of COVID-19. Ethical approval for the study was given by the Research Ethics Committee Wales, IRAS: 96194 12/WA/0148.

### Participant details

425 COVID-19 patients were recruited to the study between April 2020 and December 2021 (4 patients consented but CRFs were not completed). 405 COVID-19 patients were hospitalised and 24 recruited in the community. Demographic features including age, sex and ethnicity were recorded for all individuals at recruitment.

### Clinical data collection and processing

Following informed consent, systematic detailed demographic and clinical profiling of the COVID-19 patients was undertaken including assessment of co-morbidities, dates of onset of symptoms, first swab positive and hospital admission. clinical course and outcome of hospitalisation. Comorbidity was recorded in accordance with the ISARIC-4C clinical characterisation protocol based on the Charlston comorbidity index {8} {20}. WHO criteria scoring of severity was conducted following the ‘COVID-19 Clinical Management: living guidance’ {3}. Individuals were assigned to wave 1 (Admission 1^st^ March 2020-31^st^ August 2020), 2 (1^st^ September 2020-28^th^ April 2021 or 3 (29^th^ April 2021-June 2022) of the COVID-19 pandemic based on the date of onset of symptoms {21}. This aligned to dominant SARS-CoV-2 variants: using UK government surveillance data {22}, wild type (WT) variant dominated in wave 1 (April 2020-December 2020), alpha (B1.1.7) variant in wave 2 (20^th^ December 2020-28^th^ April 2021) and delta (B1.617) variants in wave 3 (21^st^ April 2021-14^th^ June 2022) of the COVID-19 pandemic. 72 patients had received vaccination prior to admission (‘pre-vaccinated’) of whom 2 were in the Oxford Vaccine trial {23}. 49 had received 2 vaccination doses. The median time between 1^st^ vaccination dose and symptom onset was 161 days (124-85, 95% CI) and between 2nd vaccination dose and symptom onset was 108 days (82-123, 95%CI).

Following discharge, patients were invited to follow-up visits at 3-6 and 12 months to monitor for recovery from COVID-19. This was assessed by healthcare worker delivered questionnaires that the patient completed. Fields included persistent COVID-19 related symptoms subclassified as neurological, cardiac, gastrointestinal, and COVID-19 symptoms., the MRC breathless score {24}, a modified Patient Health Questionnaire (PHQ-9) {25}/Generalised Anxiety Disorder Score {26}. In addition, patients were asked at each visit to grade their symptoms of breathlessness, cough, fatigue and sleep disturbance and indicate whether it was getting better, the same or worse. Vaccination status pre-COVID-19 and in follow-up was recorded for all individuals.

Recruitment and Clinical Data were systematically recorded using the web-based platform Open Clinica {27} including use of the self-reporting portal ‘Participate’ {28}. Clinical Reference Forms (CRFs) used for COVID-19 patients at recruitment, to discharge and in follow up are provided in Supplementary File S1: 1.1-1.4.

### Data Management and Statistical Analysis

Data generated through the CRFs was collated in Excel. Graphpad prism was used to generate the figures. All statistical analyses were performed using R software {29}. For time course analysis, ‘Date of symptom onset’ was used as Day 0 (D0). Where date of symptom onset was missing or the individual was asymptomatic, (n=34, 8% of the hospitalised cohort) a value of 14 days prior to recruitment was assigned as D0 to permit inclusion in time course data analysis. This was the median date value between symptom onset and recruitment for the overall cohort (14 days (12-14 days 95%CI), n=371).

### Statistical analysis

Unless stated otherwise, all tests were two-sided, and no multiplicity correction was used.

#### Demographic profile by COVID-19 severity

In Table 1.1 and 1.2, the relationship between severity (as a 5-level factor) and each demographic factor of interest (with numbers of levels ranging from 2 to 8) was analysed using contingency tables on which Fisher’s tests of independence were performed. The p-value from the Fisher test was obtained through Monte Carlo simulation, considering 25,000 samples, as implemented in the *fisher.test* function of the R package *stats*, applied to the contingency table generated after discarding missing values. In Figure S1.1, the pattern of missingness was analysed using a heatmap that displays the missingness status through a colour code.

**Table: 1.1:**
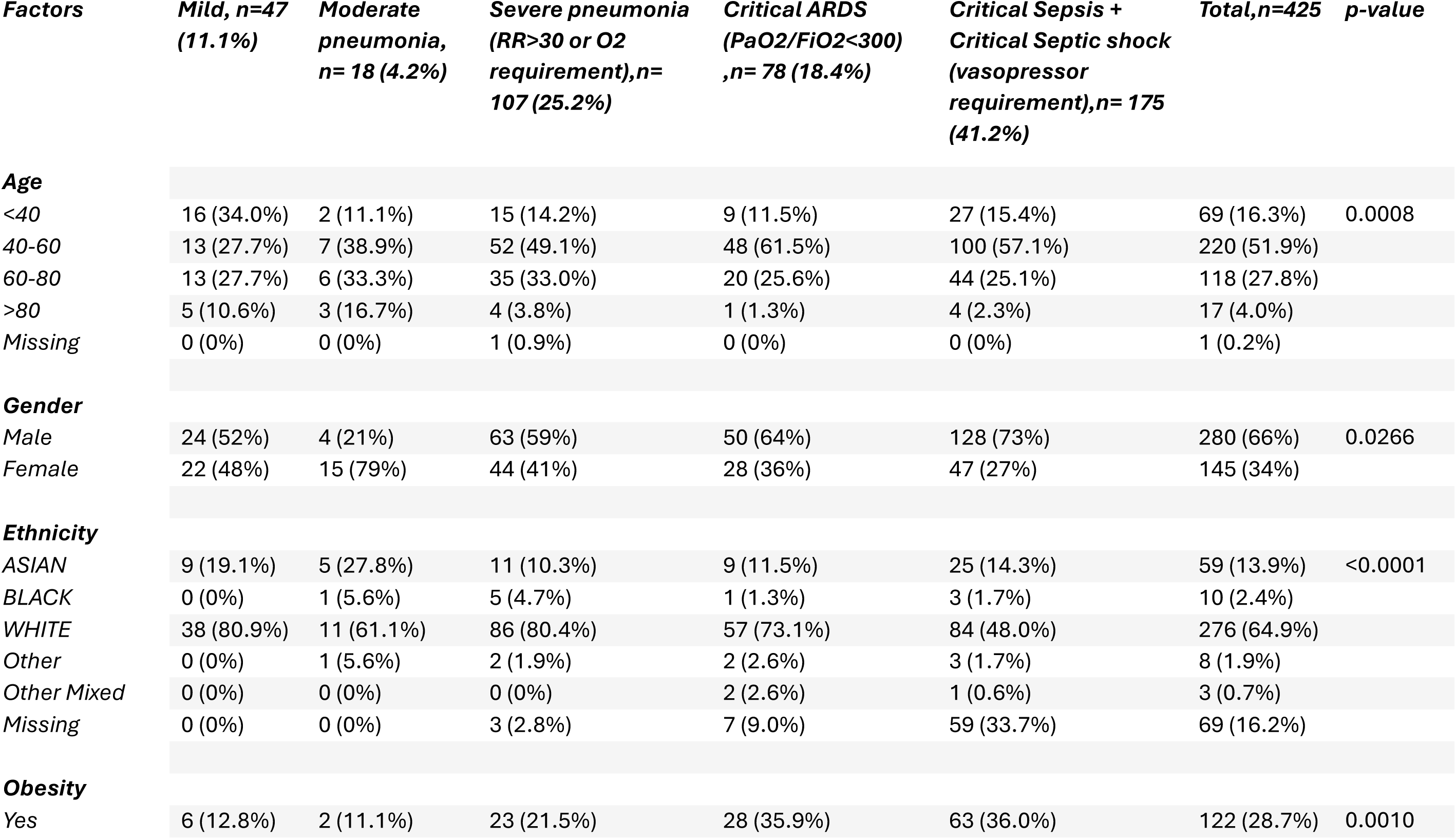
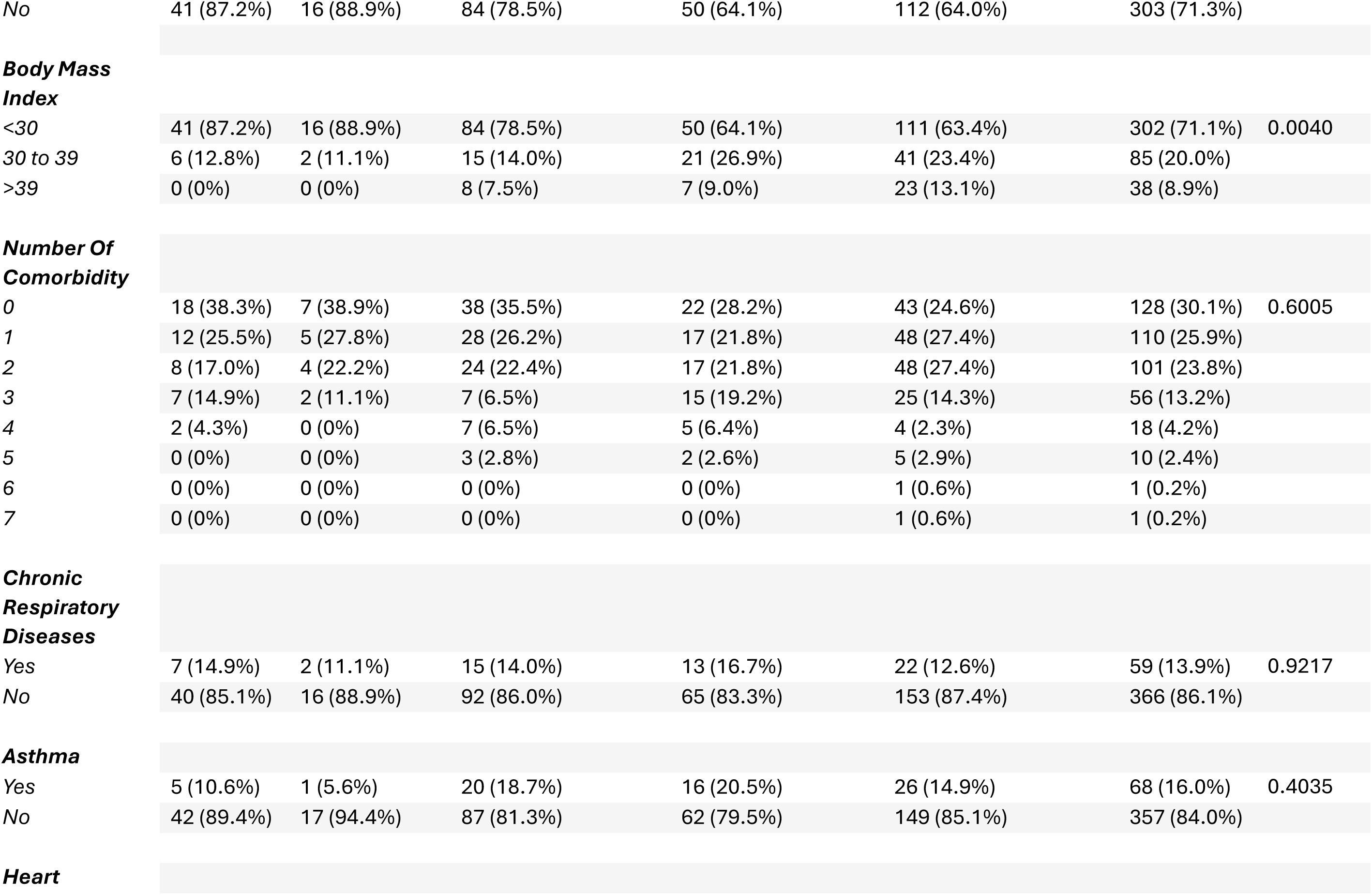
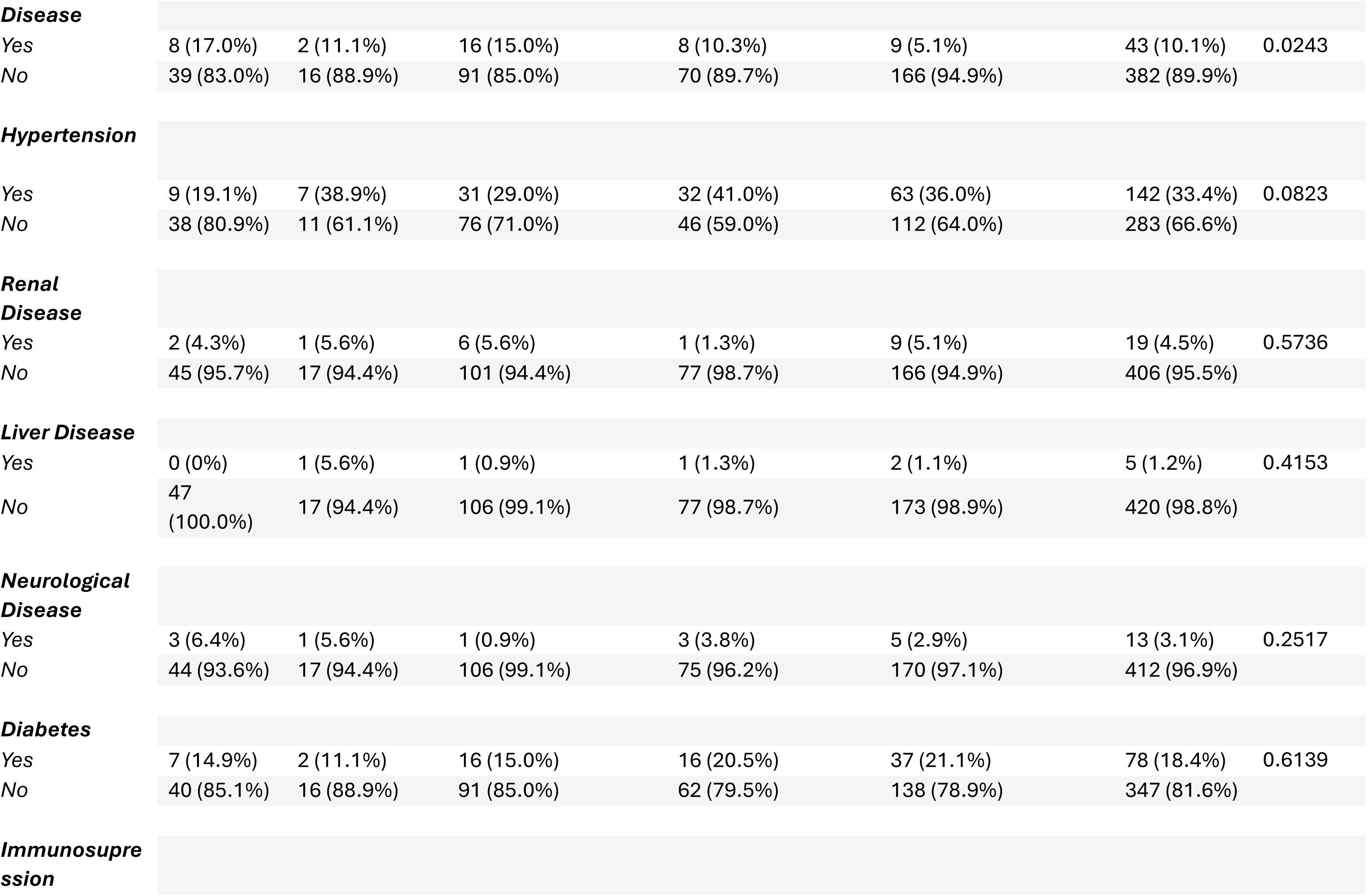
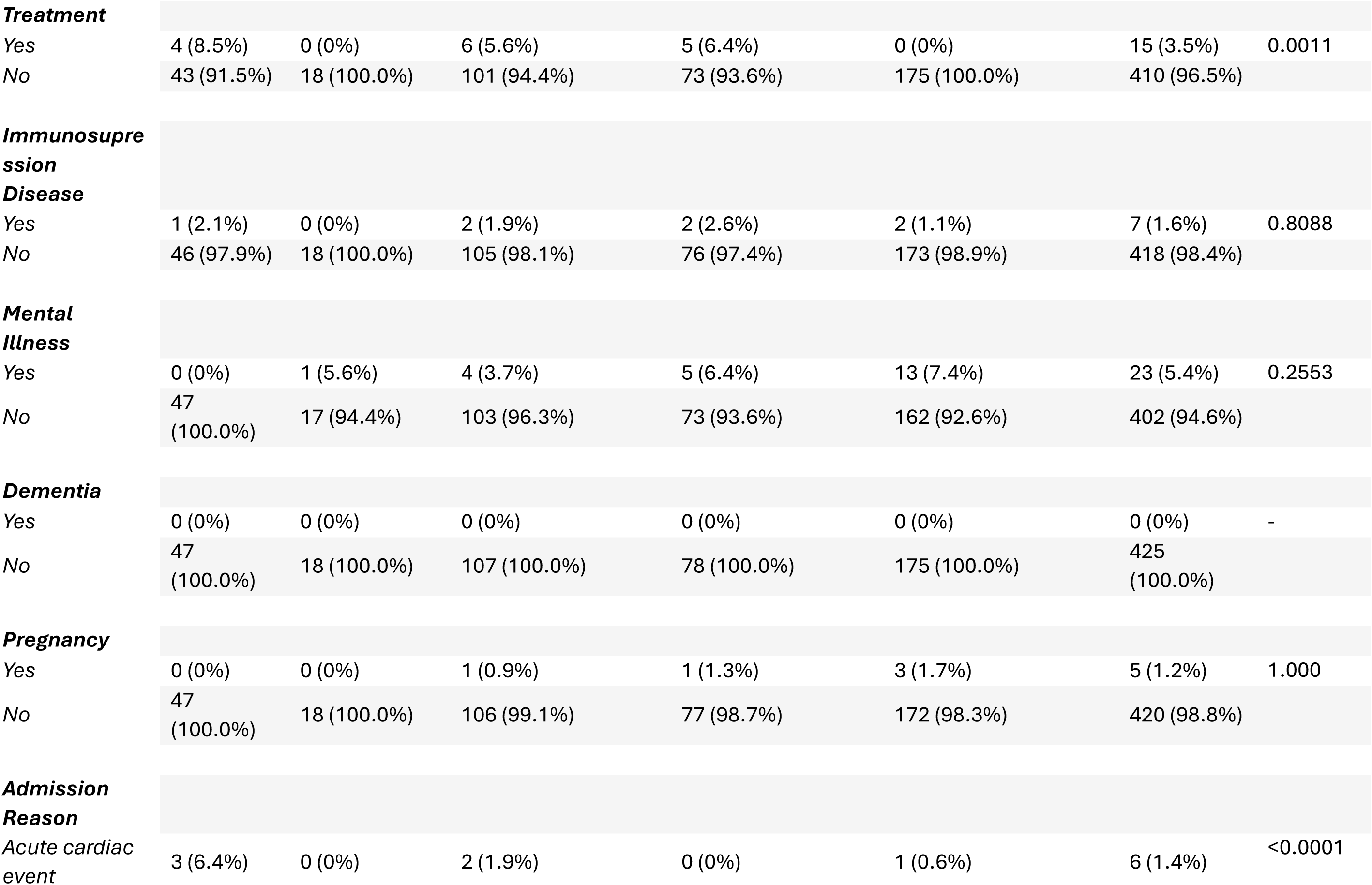
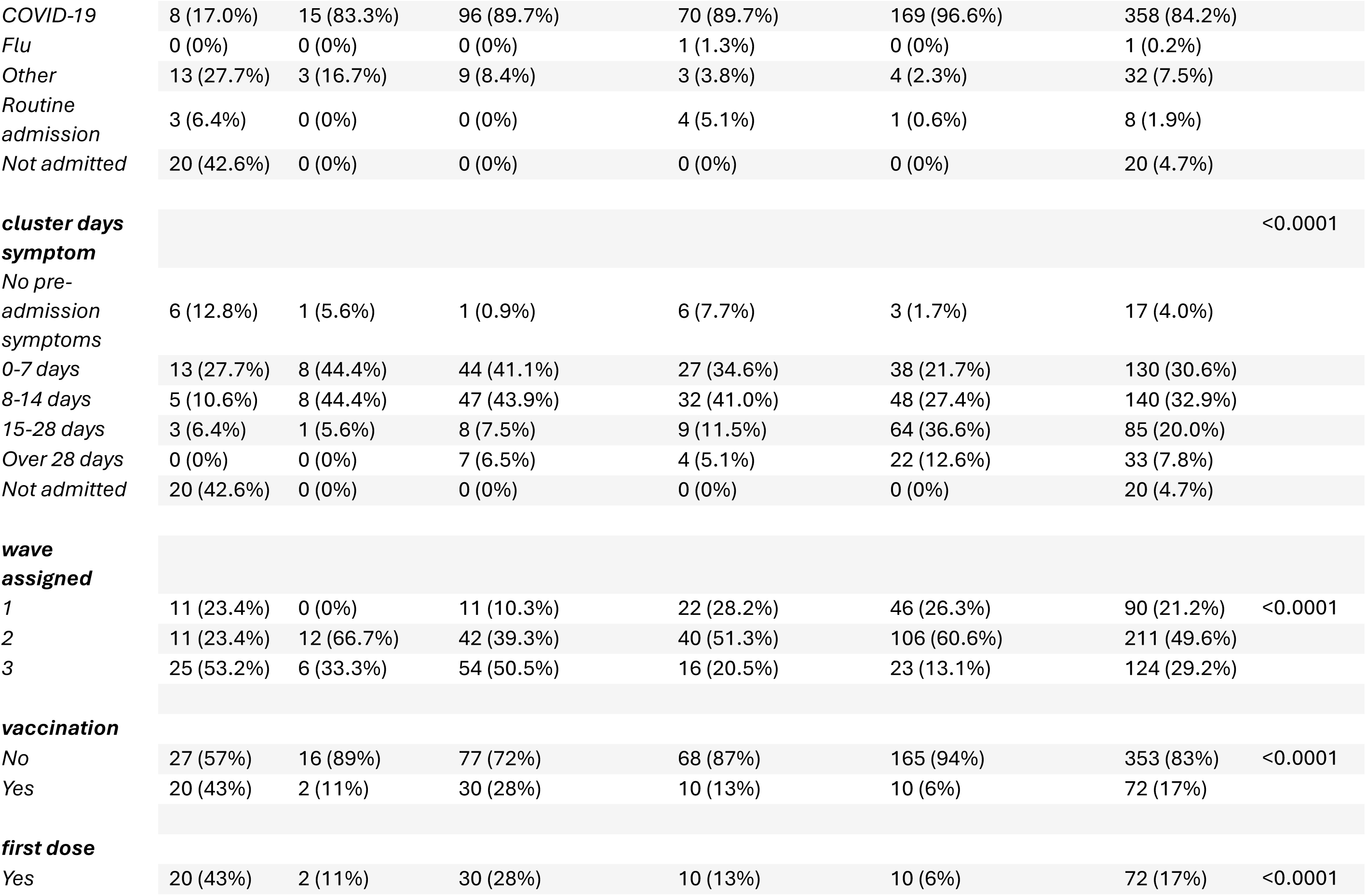
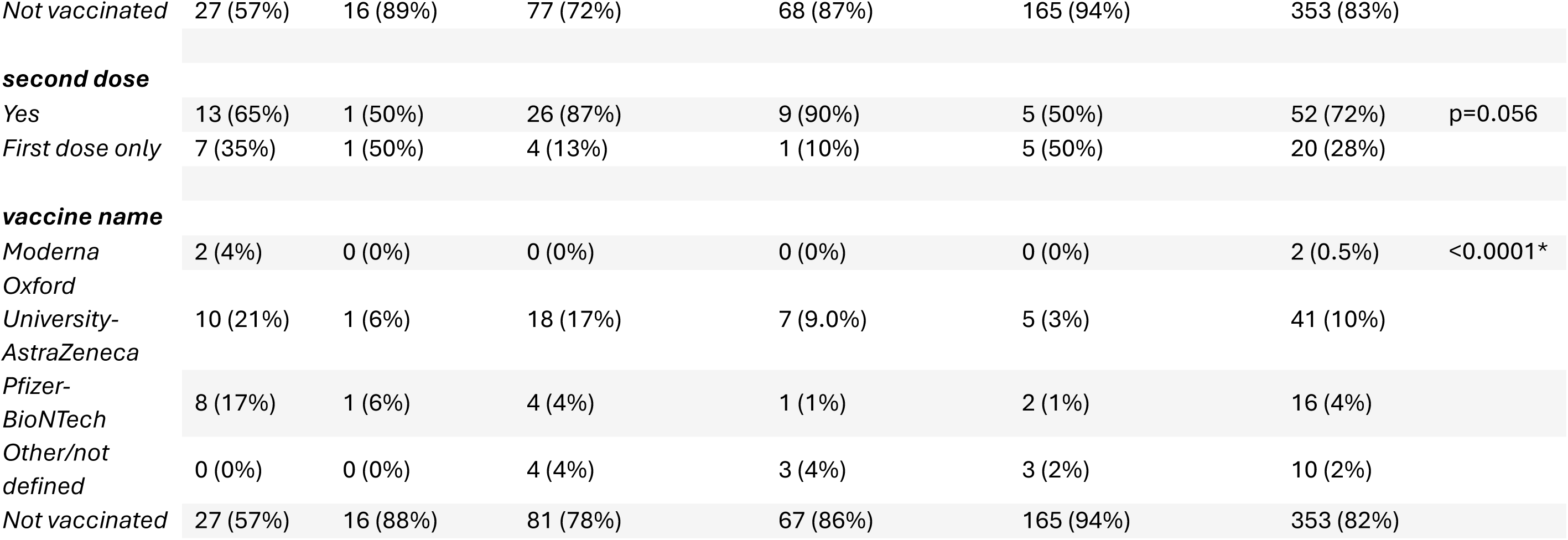
Contingency table showing the absolute number of patients for each combination of severity levels (columns) and levels of demographic factors of interest (rows). For each level of severity, the relative number of patients per level of the demographic factor of interest is also indicated as a percentage (%). The p-value from a Fisher’s test of independence between severity and each demographic factor is also provided.

**Table 1.2:** Subgroup analysis by vaccination and wave: A contingency table showing the absolute number of patients for each combination of severity levels (columns) and vaccination status (Yes/No) across each COVID wave (rows). For each level of severity, the relative number of patients per level of vaccination status is also indicated as a percentage (%). The p-value from a Fisher’s test of independence between severity and vaccination status for each wave is also provided.

| **Wave assigned** | **Vaccination** | **Mild, n=47 (11.1%)** | **Moderate pneumonia, n= 18 (4.2%)** | **Severe pneumonia (RR>30 or O2 requirement),n= 107 (25.2%)** | **Critical ARDS (PaO2/FiO2<300),n= 78 (18.4%)** | **Critical Sepsis + Critical Septic shock (vasopressor requirement),n= 175 (41.2%)** | **Total,n=425** | **P-value per wave** |
| --- | --- | --- | --- | --- | --- | --- | --- | --- |
| **1 wave** | No | 11 (100.0%) | 0 (-%) | 11 (100.0%) | 22 (100.0%) | 46 (100.0%) | 90 (100.0%) | - |
|  | Yes | 0 (0%) | 0 (-%) | 0 (0%) | 0 (0%) | 0 (0%) | 0 (0%) |  |
| **2 wave** | No | 10 (90.9%) | 11 (91.7%) | 39 (92.9%) | 39 (97.5%) | 104 (98.1%) | 203 (96.2%) | 0.1786 |
|  | Yes | 1 (9.1%) | 1 (8.3%) | 3 (7.1%) | 1 (2.5%) | 2 (1.9%) | 8 (3.8%) |  |
| **3 wave** | No | 5 (20.0%) | 4 (66.7%) | 28 (51.9%) | 6 (37.5%) | 15 (65.2%) | 58 (46.8%) | 0.0118 |
|  | Yes | 20 (80.0%) | 2 (33.3%) | 26 (48.1%) | 10 (62.5%) | 8 (34.8%) | 66 (53.2%) |  |

#### Clustering by Symptoms and Clustering by comorbidities

Symptoms and comorbidities presence/absence data were separately clustered by partitioning participants around medoids (as implemented in the *pamk* function of the R package *fpc*), based on Jaccard dissimilarity measures (obtained using the *dissimilarity* function from the R package *arules*), allowing for a suitable definition of the ‘distance’ between binary vectors. In Figure S1.2, the probability of observing each symptom and comorbidity per cluster was compared using proportion tests (as implemented in the *prop.test* function of the R package *stats*).

#### Acute clinical course of Hospitalised Patients

In Tables 3.1 to 3.6, for each clinical endpoint of interest, the presence or absence of the relevant event as a function of severity was analysed using adjusted and unadjusted logistic regression with a logit link (as fitted using the *glm* function of the R package *stats*, with the family argument set to *binomial*). Unadjusted analyses considered severity solely as an ordered categorical predictor (with *mild* as the reference group). Adjusted analyses controlled for sex, obesity, smoking status, duration of symptoms, ethnicity, vaccination status, COVID wave, age group, and symptom and comorbidity clusters. Odds ratio estimates and 95% confidence intervals, along with the corresponding Wald z-test, were calculated. Model checks, assessed using randomised quantile residuals from the R package *gamlss*, indicated a good fit of the model to the data across all clinical endpoints. In Table 3.7, survival analyses, specifically Cox regressions, are used as an alternative to the logistic regressions considered in Table 3.6. Cox regressions have the advantage of being able to handle time-to-event data and accounts for censoring, while logistic regressions are simpler, rely on different assumptions, but don’t consider time. Right-censored and left-truncated times to death were modelled using adjusted and unadjusted Cox regression analyses with a log link (as fitted using the *coxph* function of the R package *survival*). In this analysis, follow-up times for patients who were discharged or transferred were treated as right-censored, while sometimes to death were treated as left-truncated, since patients who developed symptoms and died before reaching the hospital were not recorded. Both unadjusted and adjusted analyses considered the same predictors as those used in the logistic regression. Hazard ratio estimates and 95% confidence intervals, along with the corresponding Wald z-test, were calculated. In analyses of Tables 3.1-3.7, predictors were simplified by combining levels or were removed when a strong imbalance in the number of successes and failures per combination of predictors resulted in unreasonably large standard errors due to numerical issues. This frequently occurred with the predictor severity, which was therefore treated as a 2- or 3-level factor in many analyses.

#### Long-term Recovery from COVID-19

In Tables 4.1 and 4.2, adjusted and unadjusted generalised linear mixed models for binary endpoints, as fitted using the *glmer* function from the *lme4* package with the family argument set to *binomial*, were used to analyse the effect of severity on two recovery endpoints—the MRC Breathlessness score ranged from 0-4 and was dichotomised into 0 (no breathlessness) to 1-4 (mild to severe breathlessness), and mood disorder from 0 (least disorder) to 16 based on 4 parameters (interest, feeling down, nervous and worrying) —measured at up to three time points per patient and was dichotomised into 0 vs >0. Details of the follow up questionnaires are found in Supplementary File S1.4. All models treated patients as random effects. Unadjusted analyses solely predicted the probability of success based on severity (a 3-level factor with mild as the reference group) and time (a factor with levels 3, 6, and 12 months, with 3 months as the reference group), while adjusted analyses further controlled for sex, obesity, smoking status, duration of symptoms, ethnicity, vaccination status, age group, and symptom and comorbidity clusters. Odds ratio estimates and 95% confidence intervals, along with the corresponding Wald z-test, were calculated, and p-values were not adjusted for multiplicity.

#### Trajectory of Recovery from COVID-19 to 12 months after hospital discharge

The trajectory of recovery from COVID following hospital discharge was assessed by matched pair analysis of four endpoints: MRC Breathlessness Score, Mood Disorder Score, number of COVID symptoms and Rockwood Frailty Index, collected at 3,6 and 12 months. Wilcoxon two tailed t-tests of paired data was used to test for change in symptoms over each period (Figure 4.2).

#### Persistence of COVID-19 symptoms up to 12 months by gender

The odds ratio with 95% CI of reporting persistent COVID-19 symptoms at the 12-month follow up visit by gender was determined Wald z-tests of univariate logistic regression, with p value generated for difference in symptom reporting between gender.

## Results

### Recruitment Demographic

425 patients were recruited to the study across the first 3 waves of the COVID-19 pandemic. The median age of the cohort was 53 years (95% CI: 51-56 years). 66% of patients recruited were male and of white Ethnicity. 65% of individuals recruited had at least 1 comorbidity of which hypertension dominated (33% of all individuals); 27% had 1 and 38% 2 or more comorbidities. 29% of individuals were obese of whom 1/3 had a BMI>40. 405 patients were recruited in hospital (95% of the patient cohort), of whom 88% were admitted for management of COVID-19, 2% with acute cardiac event and 10% for other reasons including acute medical review (3%) or surgical procedure (3.5%). 47% of patients were admitted to the study recruitment hospital from another hospital, <1% from a care home and 48% of patients were admitted from home (data not shown). 5% patients were recruited in the community during wave 3 of the pandemic. Using date of symptom onset to assign ‘wave’ and thus likely SARS-CoV-2 variant causing disease, 21% of patients were admitted in wave 1, 50% in wave 2 and 29% in wave 3 of the COVID-19 pandemic. Disease severity varied and was dominated by patients with severe COVID-19 disease accounting for 60% of the cohort. A detailed description of the clinical and demographic characteristics of the overall patient cohort by COVID-19 disease severity score on admission is provided in Table 1.1. We show that multiple demographic factors varied significantly by disease severity score on admission including age, gender, obesity, ethnicity, pre-morbid conditions such as heart disease and taking immunosuppressive treatment, reason for admission, duration of pre-admission symptoms, wave of the epidemic at admission, vaccination status, number of doses and vaccine type received. 17% of patients had received at least 1 vaccination at presentation, predominantly in wave 3 of the pandemic (Table 1b) of whom 27% were admitted with ARDS or critical sepsis.

### Acute Clinical Course and Outcome of hospitalised patients

The clinical course of the acute phase of COVID-19 for patients recruited in hospital (n=405) is summarised in Table 2 including the type of respiratory support given (many patients progressing through modality), clinical trial and adjunctive treatments, complications and outcome of admission. In brief, 86% of the cohort received respiratory support and over 50% of the cohort were admitted to the intensive care unit (ICU) with 22% of cohort receiving extracorporeal membrane oxygenation therapy (ECMO) (Table 1). 39% patients were recruited to clinical trials and 17% randomised to treatment arms which included convalescent plasma, biologics and antivirals. Adjunctive treatments were given to at least 24% of patients including haemofiltration, plasmapheresis and cytosorp. The median duration of ICU stay and ventilatory support by modality is shown in Figure 1.1 noting that duration of ICU stay and ventilatory support was widely spread with some patients remaining on ECMO for many months. Most of the cohort was discharged to home or for de-escalation of care to local hospital trust however 22% of patients died in hospital (Table 1) including 8 of patients (∼2% of the cohort) had been pre-vaccinated. Admission duration varied from 1 to over 128 days with Median length of stay was greatest in those who were ultimately transferred for de-escalation of care at 50 days (Figure 1.2).

**Figure 1.**
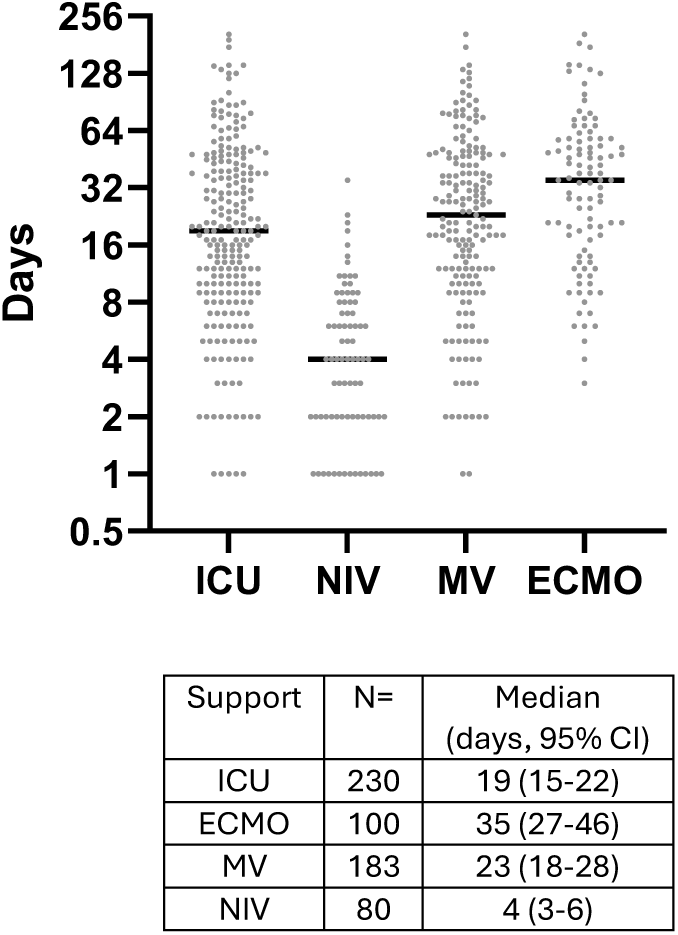
Acute clinical course of Hospitalised Patients. **Figure 1.1** Dot plot showing the number of days (y-axis, log2 scale) by clinical pathway (number of days in Intensive Care Unit (ICU) and duration of respiratory support by support modality (Non-invasive ventilation (NIV), Mechanical ventilation (MV) and Extracorporeal-membrane oxygenation (ECMO), x-axis) per patient (dots). The horizontal bar displays the median. The sample size, median estimate, and 95% confidence interval for each clinical pathway are also given.

**Figure 1.2.**
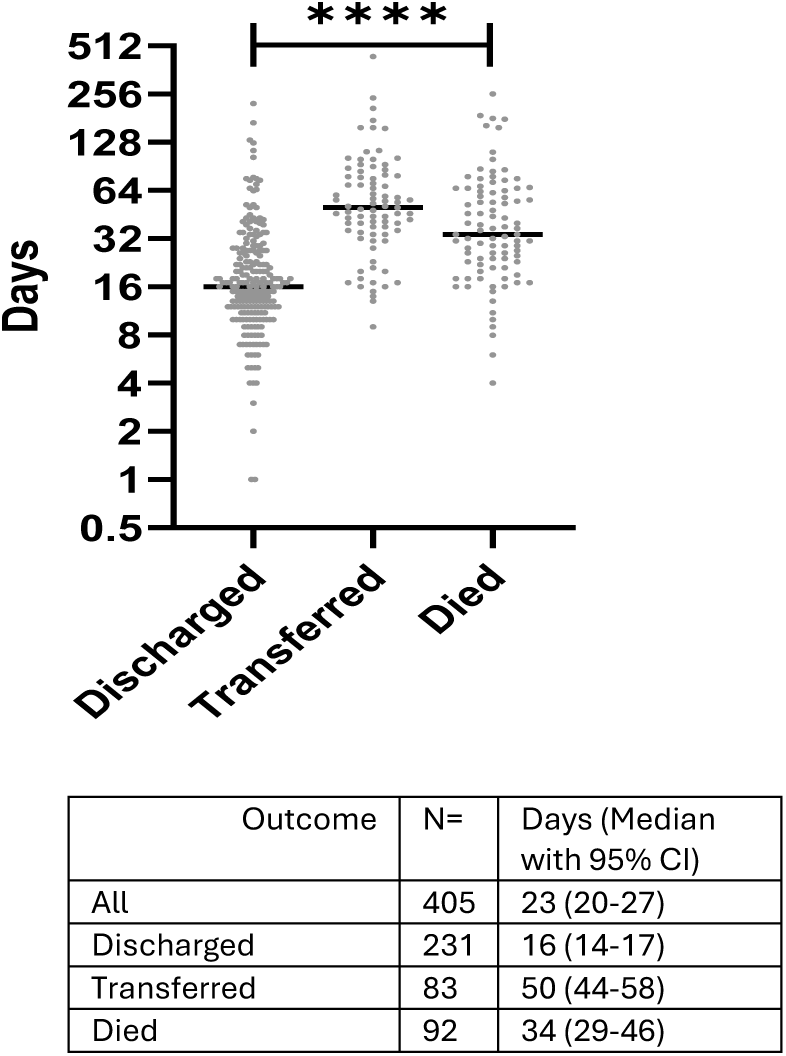
Dot plot showing the number of days from the onset of symptoms to outcome of hospital admission (y-axis, log2 scale) per patient (dots), overall and according to outcome: discharge, transfer to another hospital for de-escalation of care and death (x-axis). The horizontal bar displays the median. The sample size, median estimate, and 95% confidence interval for each outcome are indicated in the table below. The p-value of a test for equality of medians is also indicated.

**Figures 2.**
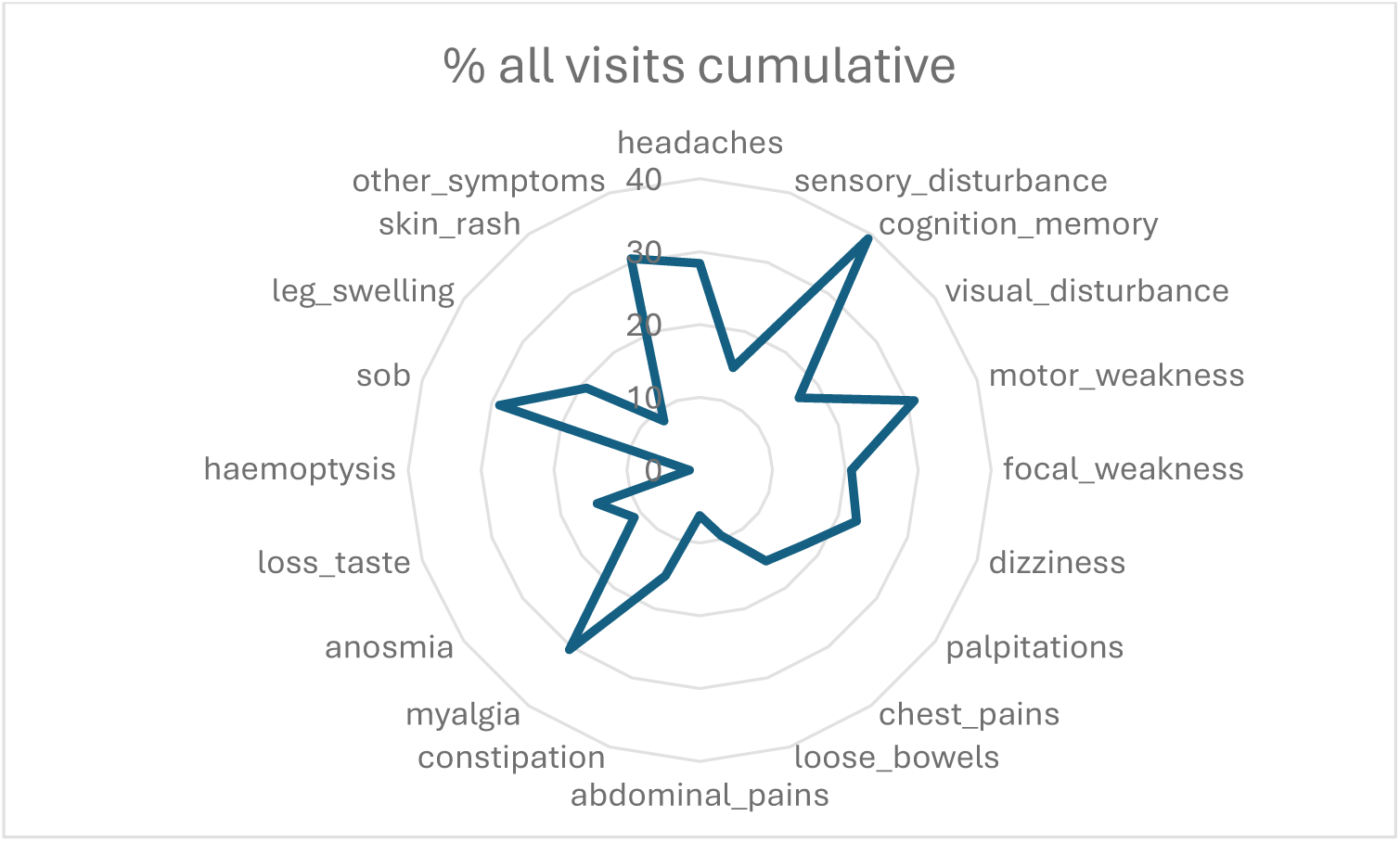
Prevalence of persisting COVID symptoms in recovery. **Figure 2.1. Symptom prevalence:** Spider plot showing the relative frequency (%) of the presence of each clinical endpoint of interest in the study cohort.

**Figure 2.2.**
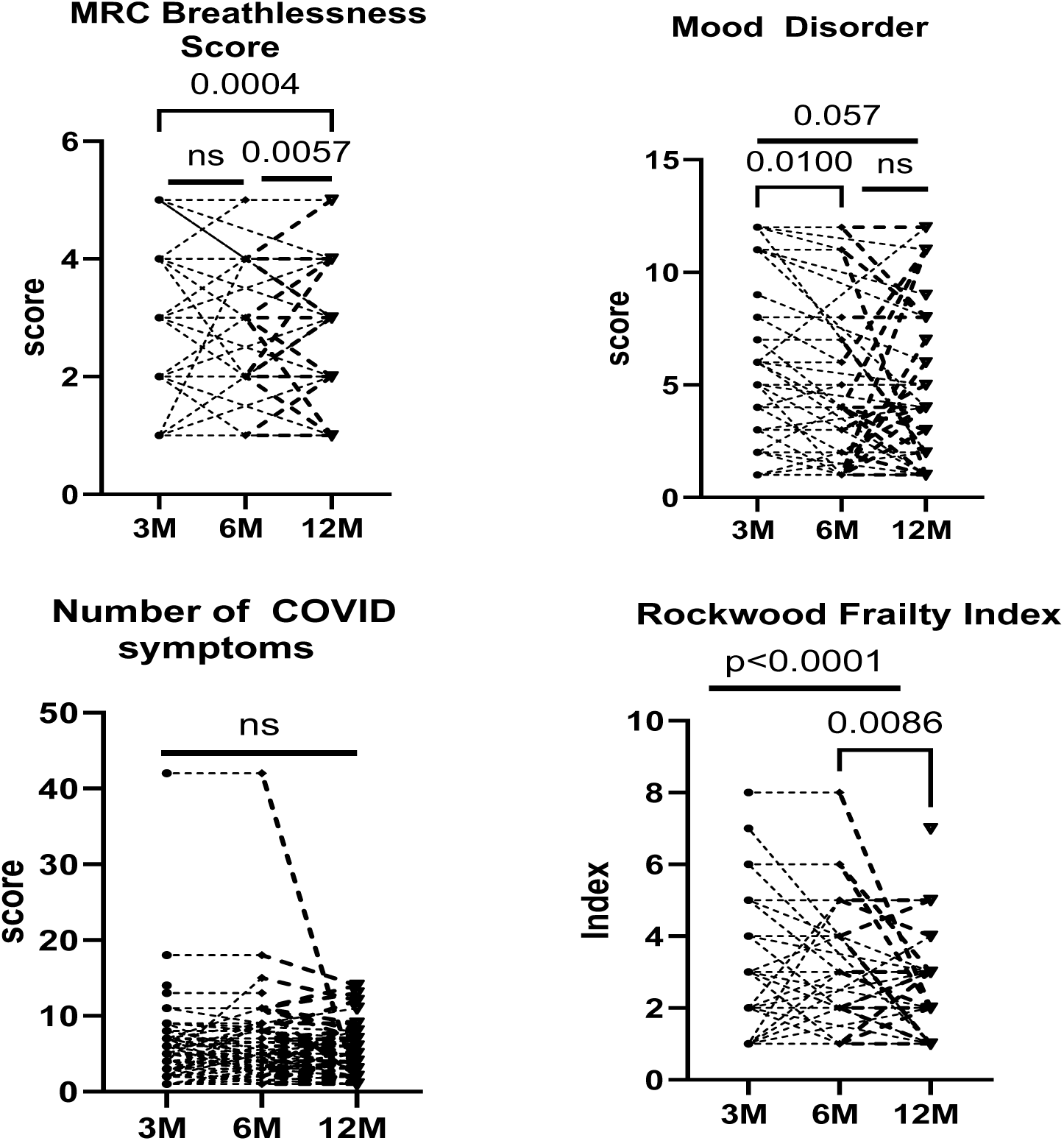
Trajectory of recovery over time: Patients’ trajectories (lines) as a function of time (x-axes) for different clinical endpoints (plots) are shown by match pair analysis (Wilcoxon 2 tailed). P-values of paired t-tests (on the linear scale) are indicated for each combination of time points.

**Figure 2.3.**
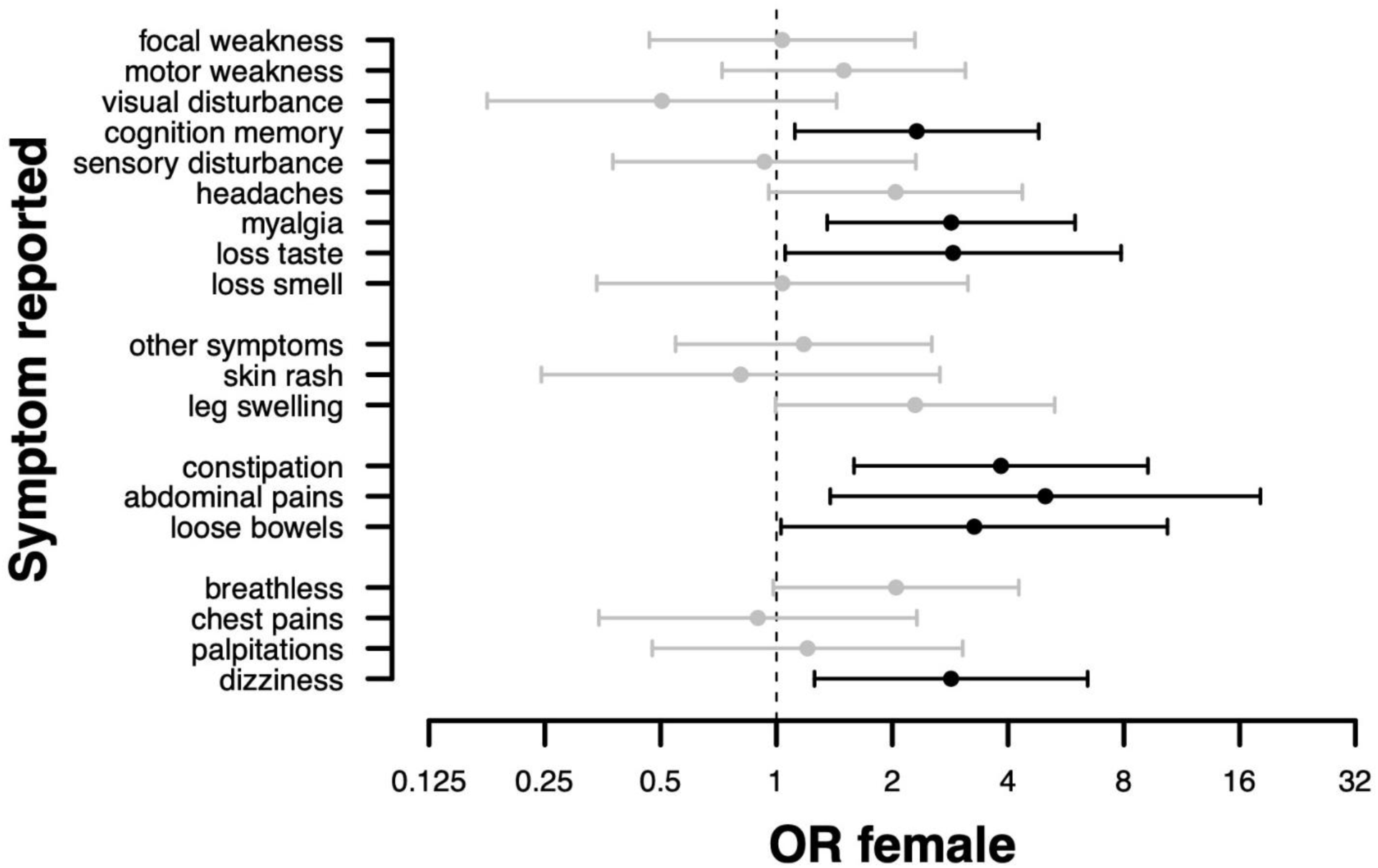
COVID-19 symptoms by gender 12 months after symptom onset. Odds ratio (OR) (x-axis) is provided related to the reporting persistent symptoms by gender (y-axis), where OR estimates and confidence intervals are respectively denoted by a dot and a segment, which are color-coded in black in grey and black; black when the odds ratio is different from 1 at the 5% type I error level*. Median time since symptom onset was 376 days (95% CI: 360-398 days). Women more likely to suffer with persistent COVID-19 symptoms: specifically, cognitive problems (p=0.024), myalgia (p=0.006), loss of taste (p=0.039), GI (abdominal pain p=0.014, constipation p=0.003, loose stool p=0.044* and dizziness (p=0.012).

**Table 2.** Clinical Course of Acute Admission and Outcome of hospitalised patients. . Number of patients requiring intensive care unit (ICU) admission, respiratory support including mode of support, enrolment in treatment trials and treatments given, complications of admission, and outcome of admission are given. Non-invasive ventilation (NIV); Mechanical ventilation (MV); Extracorporeal Membrane Oxygenation (ECMO).

| Complications |  | Medical Management | N= | % |
| --- | --- | --- | --- | --- |
| **Medical Support** |  | **ICU Admission** | 232 | 54.6 |
| **Respiratory support** |  | **Respiratory support** | 365 | 85.9 |
|  |  | Oxygen via cannula or mask | 99 | 23.3 |
|  |  | High flow nasal oxygen | 28 | 6.6 |
|  |  | NIV | 41 | 9.6 |
|  |  | MV | 86 | 20.2 |
|  |  | ECMO | 89 | 20.9 |
| **Treatment Trial** |  | **Trial treatment enrolled** | 158 | 37.2 |
|  |  | Treatment arm | 70 | 16.5 |
|  |  | Standard of care | 88 | 20.7 |
|  |  | Convalescent plasma | 11 | 2.6 |
|  |  | Convalescent plasma and colchicine | 3 | 0.7 |
|  |  | Convalescent plasma and aspirin and colchicine | 2 | 0.5 |
|  |  | Convalescent plasma and aspirin and simvastatin | 1 | 0.2 |
|  |  | Regeneron | 3 | 0.7 |
|  | mAb antiviral | Regeneron and colchicine | 1 | 0.2 |
|  |  | Regeneron and aspirin | 5 | 1.2 |
|  |  | Regeneron and Baricitinib | 1 | 0.2 |
|  |  | Baricitinib | 6 | 1.4 |
|  | mAb anti-inflammatory | Tocilizumab | 3 | 0.7 |
|  |  | Tocilizumab and colchicine | 2 | 0.5 |
|  |  | Tocilizumab and hydroxychloroquine | 2 | 0.5 |
|  |  | Rovelizumab | 1 | 0.2 |
|  |  | aspirin | 4 | 0.9 |
|  | other | aspirin and colchicine | 3 | 0.7 |
|  |  | colchicine | 1 | 0.2 |
|  |  | azithromycin | 1 | 0.2 |
|  |  | hydroxychloroquine | 11 | 2.6 |
|  |  | dexamethasone | 13 | 3.1 |
|  | steroid | dexamethasone and remdesivir | 1 | 0.2 |
|  | antiviral | lopinavir ritonavir | 1 | 0.2 |
|  | At discharge | HEAL-COVID | 4 | 0.9 |
| **Adjunctive treatment** |  | haemofiltration | 103 | 24.2 |
|  |  | plasmapheresis | 16 | 3.8 |
|  |  | CytoSorb | 23 | 5.4 |
| **Complications** |  | thrombotic events | 76 | 17.9 |
|  |  | cardiac events | 36 | 8.5 |
|  |  | neurological events | 40 | 9.4 |
|  |  | renal complications | 71 | 16.7 |
|  |  | pneumothorax | 24 | 5.6 |
|  |  | secondary bacterial pneumonia | 25 | 5.9 |
| **Outcome** |  | Discharge to Home | 230 | 54.1 |
|  |  | Transfer for de-escalation of care | 83 | 19.5 |
|  |  | Death | 92 | 21.6 |

Thrombotic, neurological, renal, cardiac events, and pneumothorax were reported in several patients of which thrombosis and renal failure dominated reported in 18% and 17% of patients respectively (Table 2). These complications were mainly seen in patients with high disease severity score. The Odds Ratio (OR) of these complications (except for cardiac event, Table 3.4) increased in patients with critical sepsis, (Severity Score 5 and 6), (Table T3.1-3.5 respectively). compared to a reference group, where the reference group varies depending on the complication. Certain demographic factors modified the point estimate of the complication risk although few reached significance. The relationship between the 11 most common symptoms of COVID and complications of admission was explored with symptoms first clustered into 2 main groups (Figure S1.2) and then evaluated to determine whether cluster membership may be an independent predictor of acute clinical course additional to COVID severity score on admission. Symptom cluster 2, a respiratory dominated symptom cluster, associated with increased risk of thrombosis and renal complications (Tables 3.1 and 3.3); admission in wave 1 (compared to wave 2 but not wave 3) with increased risk of renal complications; and symptoms for less than 1 week before admission with increased risk of pneumothorax (Table 3.5). COVID-19 severity score on admission remained a significant risk factor for acute complications of disease, except for cardiac events.

**Table 3.1-3.7:**
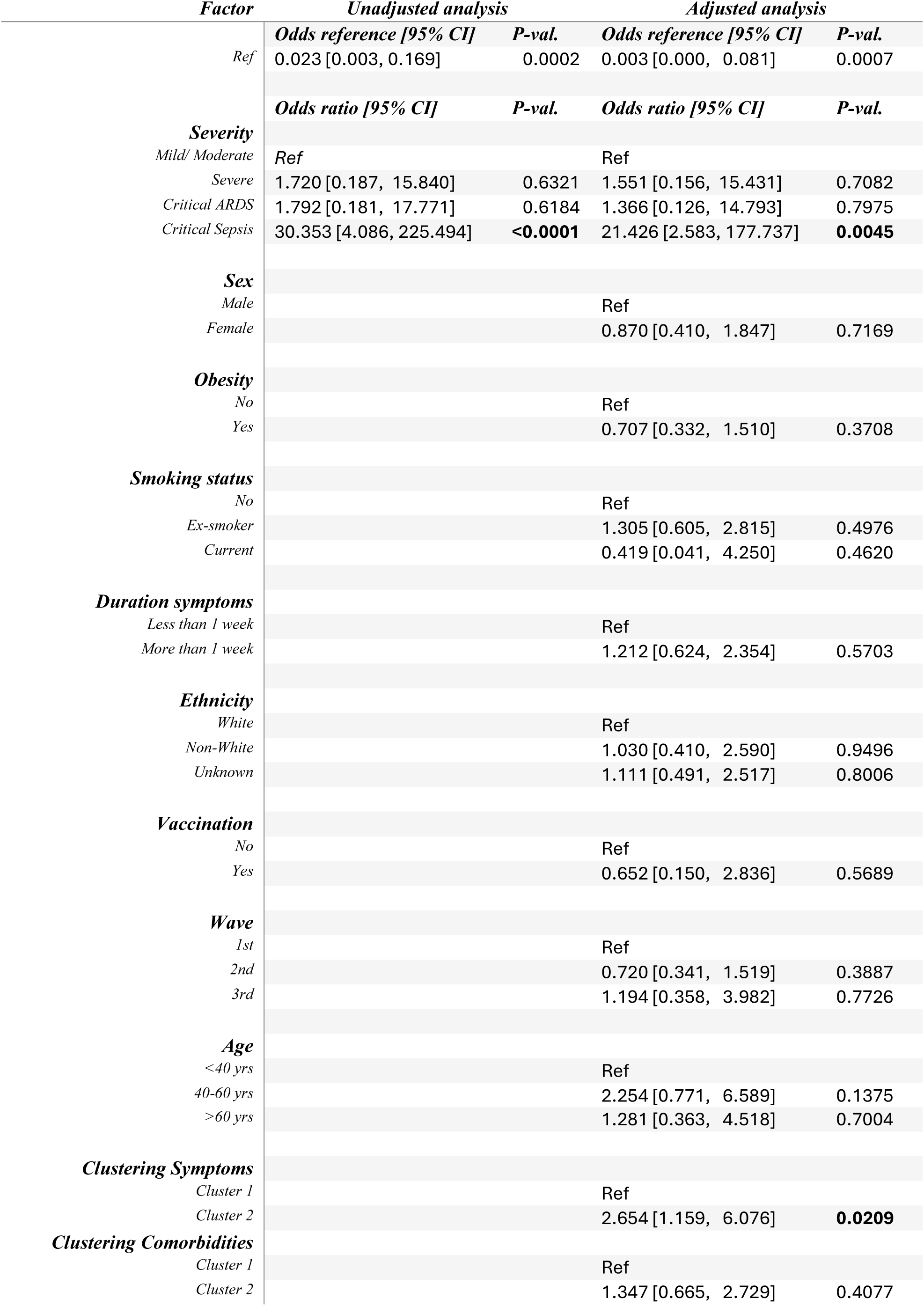
Pre-admission COVID severity score demographic factors associating with acute clinical course of hospitalised patients. **Table 3.1: Thrombotic events** Odds ratio estimates and 95% confidence intervals, along with the raw p-value from the corresponding Wald z-test, are presented for both unadjusted (left) and adjusted (right) logistic regression. These analyses aim to predict the presence or absence of the clinical endpoint of interest as a function of severity alone (unadjusted analysis) or as a function of severity plus additional potential confounders (adjusted analysis).

| *Factor* | *Unadjusted analysis* |  | *Adjusted analysis* |  |
| --- | --- | --- | --- | --- |
|  | *Odds reference [95% CI]* | *P-val.* | *Odds reference [95% CI]* | *P-val.* |
| *Ref* | 0.023 [0.003, 0.169] | 0.0002 | 0.003 [0.000, 0.081] | 0.0007 |
|  | *Odds ratio [95% CI]* | *P-val.* | *Odds ratio [95% CI]* | *P-val.* |
| **Severity** |  |  |  |  |
| *Mild/ Moderate* | *Ref* |  | *Ref* |  |
| *Severe* | 1.720 [0.187, 15.840] | 0.6321 | 1.551 [0.156, 15.431] | 0.7082 |
| *Critical ARDS* | 1.792 [0.181, 17.771] | 0.6184 | 1.366 [0.126, 14.793] | 0.7975 |
| *Critical Sepsis* | 30.353 [4.086, 225.494] | **<0.0001** | 21.426 [2.583, 177.737] | **0.0045** |
| **Sex** |  |  |  |  |
| *Male* |  |  | *Ref* |  |
| *Female* |  |  | 0.870 [0.410, 1.847] | 0.7169 |
| **Obesity** |  |  |  |  |
| *No* |  |  | *Ref* |  |
| *Yes* |  |  | 0.707 [0.332, 1.510] | 0.3708 |
| **Smoking status** |  |  |  |  |
| *No* |  |  | *Ref* |  |
| *Ex-smoker* |  |  | 1.305 [0.605, 2.815] | 0.4976 |
| *Current* |  |  | 0.419 [0.041, 4.250] | 0.4620 |
| **Duration symptoms** |  |  |  |  |
| *Less than 1 week* |  |  | *Ref* |  |
| *More than 1 week* |  |  | 1.212 [0.624, 2.354] | 0.5703 |
| **Ethnicity** |  |  |  |  |
| *White* |  |  | *Ref* |  |
| *Non-White* |  |  | 1.030 [0.410, 2.590] | 0.9496 |
| *Unknown* |  |  | 1.111 [0.491, 2.517] | 0.8006 |
| **Vaccination** |  |  |  |  |
| *No* |  |  | *Ref* |  |
| *Yes* |  |  | 0.652 [0.150, 2.836] | 0.5689 |
| **Wave** |  |  |  |  |
| *1st* |  |  | *Ref* |  |
| *2nd* |  |  | 0.720 [0.341, 1.519] | 0.3887 |
| *3rd* |  |  | 1.194 [0.358, 3.982] | 0.7726 |
| **Age** |  |  |  |  |
| *<40 yrs* |  |  | *Ref* |  |
| *40-60 yrs* |  |  | 2.254 [0.771, 6.589] | 0.1375 |
| *>60 yrs* |  |  | 1.281 [0.363, 4.518] | 0.7004 |
| **Clustering Symptoms** |  |  |  |  |
| *Cluster 1* |  |  | *Ref* |  |
| *Cluster 2* |  |  | 2.654 [1.159, 6.076] | **0.0209** |
| **Clustering Comorbidities** |  |  |  |  |
| *Cluster 1* |  |  | *Ref* |  |
| *Cluster 2* |  |  | 1.347 [0.665, 2.729] | 0.4077 |

**Table 3.2:**
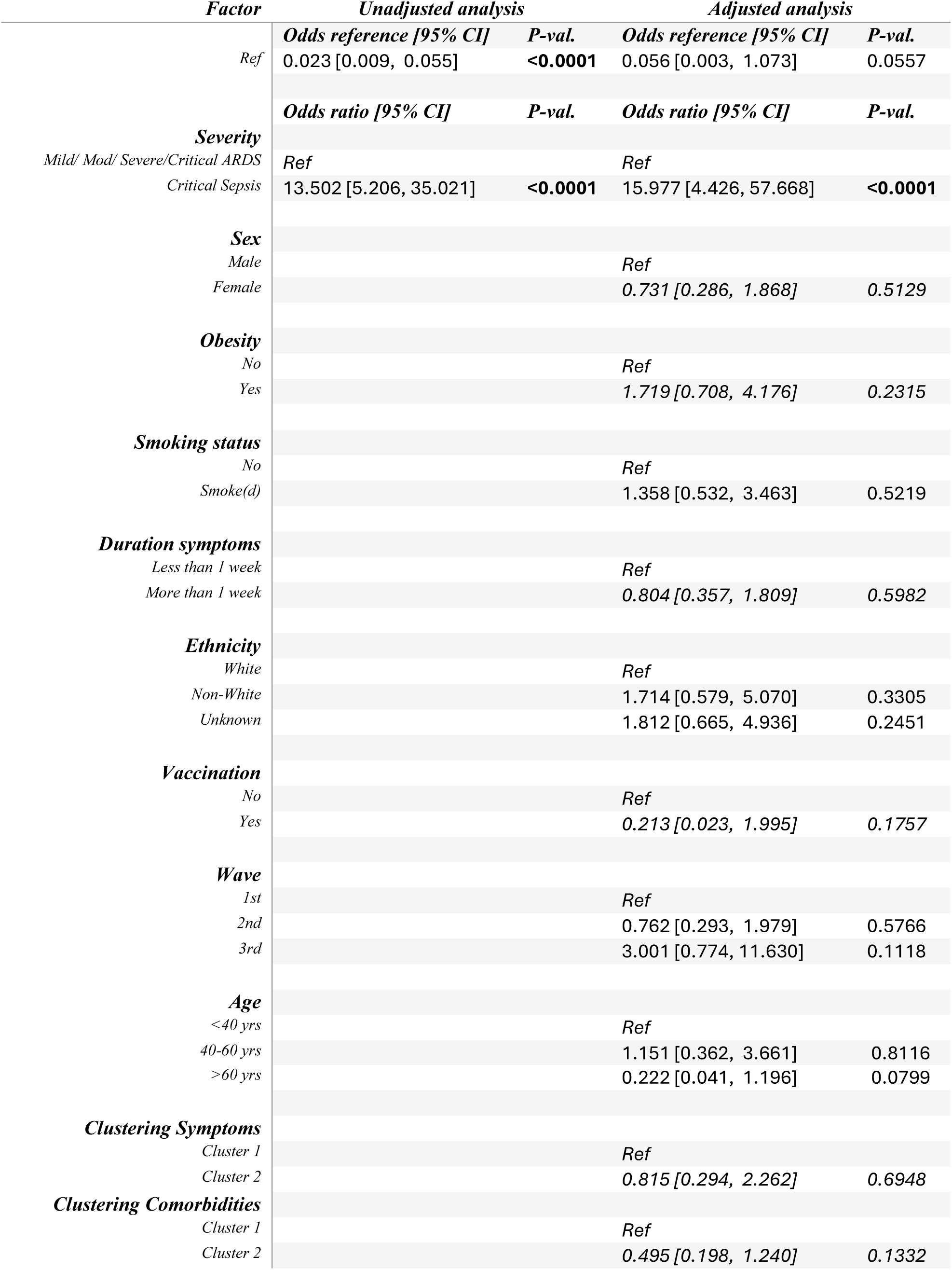
Neurological events. Odds ratio estimates and 95% confidence intervals, along with the raw p-value from the corresponding Wald z-test, are presented for both unadjusted (left) and adjusted (right) logistic regression. These analyses aim to predict the presence or absence of the clinical endpoint of interest as a function of severity alone (unadjusted analysis) or as a function of severity plus additional potential confounders (adjusted analysis).

**Table 3.3:**
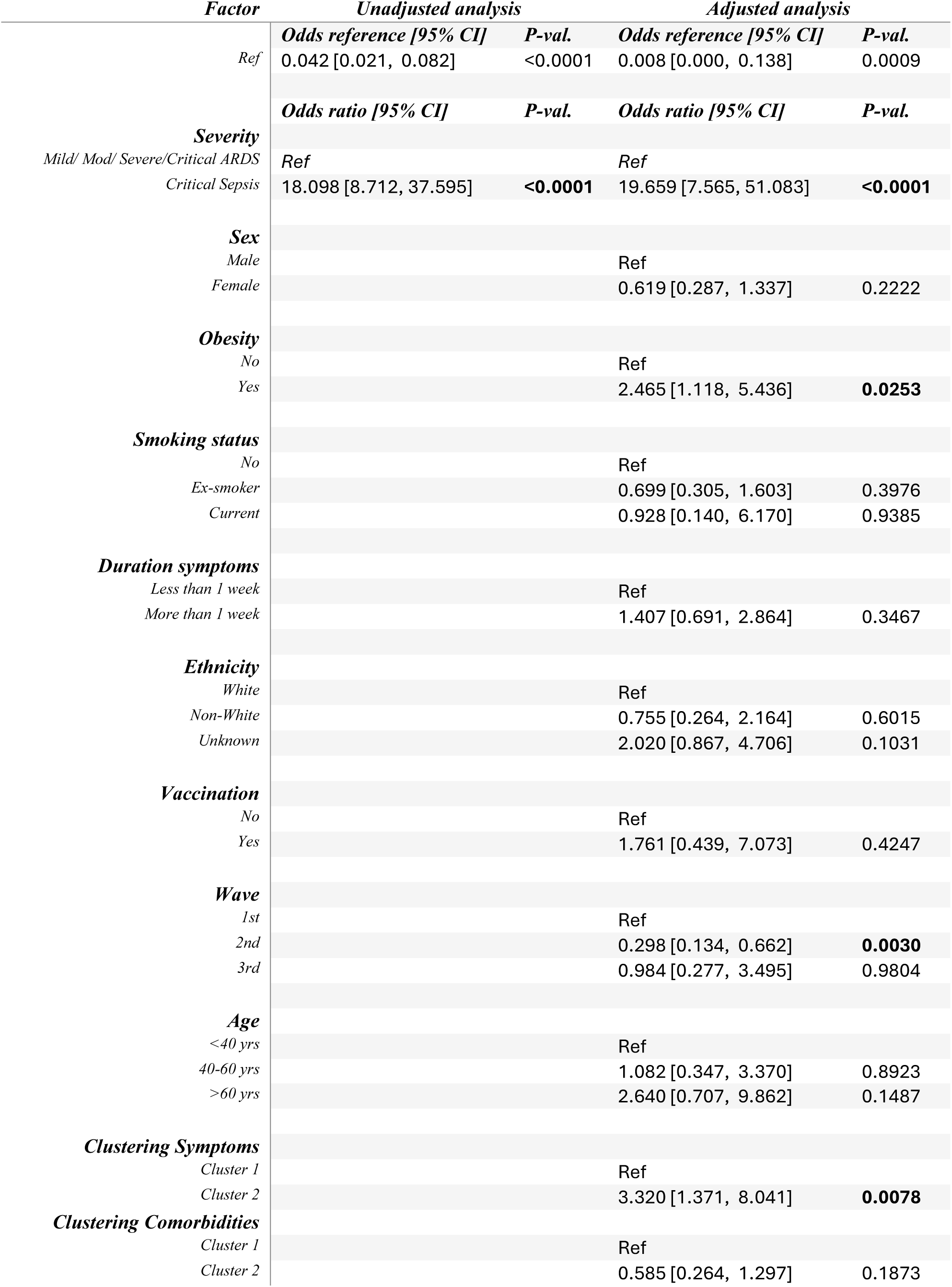
Renal complications. Odds ratio estimates and 95% confidence intervals, along with the raw p-value from the corresponding Wald z-test, are presented for both unadjusted (left) and adjusted (right) logistic regression. These analyses aim to predict the presence or absence of the clinical endpoint of interest as a function of severity alone (unadjusted analysis) or as a function of severity plus additional potential confounders (adjusted analysis).

**Table 3.4:** Cardiac events. Odds ratio estimates and 95% confidence intervals, along with the raw p-value from the corresponding Wald z-test, are presented for both unadjusted (left) and adjusted (right) logistic regression. These analyses aim to predict the presence or absence of the clinical endpoint of interest as a function of severity alone (unadjusted analysis) or as a function of severity plus additional potential confounders (adjusted analysis).

| *Factor* | *Unadjusted analysis* |  | *Adjusted analysis* |  |
| --- | --- | --- | --- | --- |
|  | *Odds reference [95% CI]* | *P-val.* | *Odds reference [95% CI]* | *P-val.* |
| *Ref* | 0.073 [0.023, 0.236] | <0.0001 | 0.003 [0.000, 0.140] | 0.0027 |
|  | *Odds ratio [95% CI]* | *P-val.* | *Odds ratio [95% CI]* | *P-val.* |
| **Severity** |  |  |  |  |
| *Mild/ Moderate* | Ref |  | Ref |  |
| *Severe* | 0.406 [0.079, 2.095] | 0.2816 | 1.130 [0.105, 12.207] | 0.9196 |
| *Critical ARDS* | 0.569 [0.110, 2.952] | 0.5024 | 1.405 [0.132, 14.914] | 0.7780 |
| *Critical Sepsis* | 3.080 [0.897, 10.573] | 0.0739 | 6.477 [0.781, 53.715] | 0.0834 |
| **Sex** |  |  |  |  |
| *Male* |  |  | Ref |  |
| *Female* |  |  | 0.992 [0.417, 2.358] | 0.9856 |
| **Obesity** |  |  |  |  |
| *No* |  |  | Ref |  |
| *Yes* |  |  | 1.468 [0.622, 3.465] | 0.3806 |
| **Smoking status** |  |  |  |  |
| *No* |  |  | Ref |  |
| *Ex-smoker* |  |  | 0.652 [0.241, 1.769] | 0.4013 |
| *Current* |  |  | 1.062 [0.114, 9.915] | 0.9577 |
| **Duration symptoms** |  |  |  |  |
| *Less than 1 week* |  |  | Ref |  |
| *More than 1 week* |  |  | 0.657 [0.298, 1.449] | 0.2983 |
| **Ethnicity** |  |  |  |  |
| *White* |  |  | Ref |  |
| *Non-White* |  |  | 1.065 [0.345, 3.283] | 0.9133 |
| *Unknown* |  |  | 1.036 [0.376, 2.857] | 0.9449 |
| **Vaccination** |  |  |  |  |
| *No* |  |  | Ref |  |
| *Yes* |  |  | 2.944 [0.630, 13.769] | 0.1701 |
| **Wave** |  |  |  |  |
| *1st* |  |  | Ref |  |
| *2nd* |  |  | 0.854 [0.342, 2.132] | 0.7347 |
| *3rd* |  |  | 0.562 [0.122, 2.598] | 0.4611 |
| **Age** |  |  |  |  |
| *<40 yrs* |  |  | Ref |  |
| *40-60 yrs* |  |  | 1.335 [0.372, 4.793] | 0.6575 |
| *>60 yrs* |  |  | 1.585 [0.362, 6.938] | 0.5407 |
| **Clustering Symptoms** |  |  |  |  |
| *Cluster 1* |  |  | Ref |  |
| *Cluster 2* |  |  | 2.006 [0.719, 5.601] | 0.1837 |
| **Clustering Comorbidities** |  |  |  |  |
| *Cluster 1* |  |  | Ref |  |
| *Cluster 2* |  |  | 1.969 [0.851, 4.558] | 0.1136 |

**Table 3.5:** Pneumothorax. Odds ratio estimates and 95% confidence intervals, along with the raw p-value from the corresponding Wald z-test, are presented for both unadjusted (left) and adjusted (right) logistic regression. These analyses aim to predict the presence or absence of the clinical endpoint of interest as a function of severity alone (unadjusted analysis) or as a function of severity plus additional potential confounders (adjusted analysis).

| *Factor* | *Unadjusted analysis* |  | *Adjusted analysis* |  |
| --- | --- | --- | --- | --- |
|  | *Odds reference [95% CI]* | *P-val.* | *Odds reference [95% CI]* | *P-val.* |
| *Ref* | 0.023 [0.009, 0.055] | <0.0001 | 0.005 [0.000, 0.181] | 0.0040 |
|  | *Odds ratio [95% CI]* | *P-val.* | *Odds ratio [95% CI]* | *P-val.* |
| **Severity** |  |  |  |  |
| *Mild/ Mod/ Severe/Critical ARDS* | Ref |  | Ref |  |
| *Critical Sepsis* | 6.339 [2.349, 17.109] | **0.0003** | 4.248 [1.224, 14.751] | **0.0227** |
| **Sex** |  |  |  |  |
| *Male* |  |  | Ref |  |
| *Female* |  |  | 0.946 [0.331, 2.703] | 0.9167 |
| **Obesity** |  |  |  |  |
| *No* |  |  | Ref |  |
| *Yes* |  |  | 0.773 [0.267, 2.241] | 0.6353 |
| **Smoking status** |  |  |  |  |
| *No* |  |  | Ref |  |
| *Smoke(d)* |  |  | 0.824 [0.268, 2.535] | 0.7360 |
| **Duration symptoms** |  |  |  |  |
| *Less than 1 week* |  |  | Ref |  |
| *More than 1 week* |  |  | 0.322 [0.120, 0.869] | **0.0252** |
| **Ethnicity** |  |  |  |  |
| *White* |  |  | Ref |  |
| *Non-White* |  |  | 0.534 [0.130, 2.192] | 0.3836 |
| *Unknown* |  |  | 1.008 [0.313, 3.254] | 0.9888 |
| **Vaccination** |  |  |  |  |
| *No* |  |  | Ref |  |
| *Yes* |  |  | 0.307 [0.031, 3.014] | 0.3110 |
| **Wave** |  |  |  |  |
| *1st* |  |  | Ref |  |
| *2nd* |  |  | 1.294 [0.400, 4.186] | 0.6670 |
| *3rd* |  |  | 3.638 [0.790, 16.762] | 0.0976 |
| **Age** |  |  |  |  |
| *<40 yrs* |  |  | Ref |  |
| *40-60 yrs* |  |  | 1.512 [0.384, 5.955] | 0.5540 |
| *>60 yrs* |  |  | 0.162 [0.014, 1.810] | 0.1394 |
| **Clustering Symptoms** |  |  |  |  |
| *Cluster 1* |  |  | Ref |  |
| *Cluster 2* |  |  | 3.258 [0.794, 13.370] | 0.1012 |
| **Clustering Comorbidities** |  |  |  |  |
| *Cluster 1* |  |  | Ref |  |
|  |  |  | 1.155 [0.425, 3.141] | 0.7775 |

**Table 3.6:** Alive at end of local treatment (logistic model) Odds ratio estimates and 95% confidence intervals, along with the raw p-value from the corresponding Wald z-test, are presented for both unadjusted (left) and adjusted (right) logistic regression. These analyses aim to predict the presence or absence of the clinical endpoint of interest as a function of severity alone (unadjusted analysis) or as a function of severity plus additional potential confounders (adjusted analysis).

| *Factor* | *Unadjusted analysis* |  | *Adjusted analysis* |  |
| --- | --- | --- | --- | --- |
|  | *Odds reference [95% CI]* | *P-val.* | *Odds reference [95% CI]* | *P-val.* |
| *Ref* | 20.571 [9.635, 43.922] | <0.0001 | 88.814 [5.408, 1458.586] | 0.0017 |
|  | *Odds ratio [95% CI]* | *P-val.* | *Odds ratio [95% CI]* | *P-val.* |
| **Severity** |  |  |  |  |
| *Mild/ Moderate* | *Ref* |  | *Ref* |  |
| *Critical ARDS* | 0.425 [0.148, 1.220] | 0.1118 | 0.334 [0.088, 1.266] | 0.1068 |
| *Critical Sepsis* | 0.063 [0.028, 0.142] | <0.0001 | 0.040 [0.012, 0.134] | <0.0001 |
| **Sex** |  |  |  |  |
| *Male* |  |  | *Ref* |  |
| *Female* |  |  | 1.220 [0.568, 2.617] | 0.6104 |
| **Obesity** |  |  |  |  |
| *No* |  |  | *Ref* |  |
| *Yes* |  |  | 0.958 [0.454, 2.018] | 0.9095 |
| **Smoking status** |  |  |  |  |
| *Never smoked* |  |  | *Ref* |  |
| *Smoke(d)* |  |  | 1.264 [0.587, 2.722] | 0.5493 |
| **Duration symptoms** |  |  |  |  |
| *Less than 1 week* |  |  | *Ref* |  |
| *More than 1 week* |  |  | 0.786 [0.393, 1.570] | 0.4948 |
| **Ethnicity** |  |  |  |  |
| *White* |  |  | *Ref* |  |
| *Non-White* |  |  | 1.283 [0.467, 3.525] | 0.6290 |
| *Unknown* |  |  | 0.152 [0.066, 0.353] | <0.0001 |
| **Vaccination** |  |  |  |  |
| *No* |  |  | *Ref* |  |
| *Yes* |  |  | 0.853 [0.225, 3.244] | 0.8160 |
| **Wave** |  |  |  |  |
| *1st* |  |  | *Ref* |  |
| *2nd* |  |  | 0.887 [0.396, 1.985] | 0.7699 |
| *3rd* |  |  | 0.265 [0.068, 1.027] | 0.0546 |
| **Age** |  |  |  |  |
| *<40 yrs* |  |  | *Ref* |  |
| *40-60 yrs* |  |  | 0.361 [0.102, 1.280] | 0.1146 |
| *>60 yrs* |  |  | 0.205 [0.050, 0.834] | 0.0269 |
| **Clustering Symptoms** |  |  |  |  |
| *Cluster 1* |  |  | *Ref* |  |
| *Cluster 2* |  |  | 0.774 [0.355, 1.690] | 0.5205 |
| **Clustering Comorbidities** |  |  |  |  |
| *Cluster 1* |  |  | *Ref* |  |
| *Cluster 2* |  |  | 2.679 [1.206, 5.948] | 0.0155 |

**Table 3.7:** Dead at end of local treatment (survival model) Hazard ratio estimates and 95% confidence intervals, along with the raw p-value from the corresponding Wald z-test, are presented for both unadjusted (left) and adjusted (right) Cox regression analyses. These analyses aim to assess the hazard of death as a function of severity alone (unadjusted analysis) or as a function of severity in addition to other potential confounders (adjusted analysis).

| *Factor* | *Unadjusted analysis* |  | *Adjusted analysis* |  |
| --- | --- | --- | --- | --- |
|  | *Hazard ratio [95% CI]* | *P-val.* | *Hazard ratio [95% CI]* | *P-val.* |
| **Severity** |  |  |  |  |
| *Mild/ Moderate* | Ref |  | Ref |  |
| *Critical ARDS* | 1.524 [0.378, 6.144] | 0.5536 | 3.263 [0.472, 22.546] | 0.2305 |
| *Critical Sepsis* | 2.694 [0.808, 8.985] | 0.1068 | 30.920 [3.128, 305.606] | **0.0033** |
| **Sex** |  |  |  |  |
| *Male* |  |  | Ref |  |
| *Female* |  |  | 0.384 [0.116, 1.272] | 0.1175 |
| **Obesity** |  |  |  |  |
| *No* |  |  | Ref |  |
| *Yes* |  |  | 2.013 [0.634, 6.394] | 0.2353 |
| **Smoking status** |  |  |  |  |
| *Never smoked* |  |  | Ref |  |
| *Smoke(d)* |  |  | 2.151 [0.581, 7.962] | 0.2515 |
| **Duration symptoms** |  |  |  |  |
| *Less than 1 week* |  |  | Ref |  |
| *More than 1 week* |  |  | 0.180 [0.041, 0.789] | **0.0230** |
| **Ethnicity** |  |  |  |  |
| *White* |  |  | Ref |  |
| *Non-White & Unknown* |  |  | 0.378 [0.089, 1.603] | 0.1869 |
| **Vaccination** |  |  |  |  |
| *No* |  |  | Ref |  |
| *Yes* |  |  | 6.360 [1.226, 33.007] | **0.0277** |
| **Clustering Symptoms** |  |  |  |  |
| *Cluster 1* |  |  | Ref |  |
| *Cluster 2* |  |  | 0.243 [0.044, 1.339] | 0.1043 |
| **Clustering Comorbidities** |  |  |  |  |
| *Cluster 1* |  |  | Ref |  |
| *Cluster 2* |  |  | 0.030 [0.003, 0.318] | **0.0035** |

**Table 4.1-4.2.** Clinical Recovery from COVID-19 to 12 months after admission. **Table 4.1: MRC breathlessness score** Odds ratio estimates and 95% confidence intervals, along with the raw p-value from the corresponding Wald z-test, are presented for both unadjusted (left) and adjusted (right) logistic regression. These analyses aim to predict the presence or absence of the clinical endpoint of interest as a function of severity alone (unadjusted analysis) or as a function of severity plus additional potential confounders (adjusted analysis).

| *Factor* | *Unadjusted analysis* |  | *Adjusted analysis* |  |
| --- | --- | --- | --- | --- |
|  | *Odds reference [95% CI]* | *P-val.* | *Odds reference [95% CI]* | *P-val.* |
| *Ref* | 1.581 [0.494, 5.062] | 0.4402 | 0.004 [0.000, 28.212] | 0.2224 |
|  | *Odds ratio [95% CI]* | *P-val.* | *Odds ratio [95% CI]* | *P-val.* |
| **Severity** |  |  |  |  |
| *Mild/ Moderate/ Severe* | Ref |  | Ref |  |
| *Critical ARDS* | 1.042 [0.161, 6.755] | 0.9655 | 2.819 [0.188, 42.208] | 0.4528 |
| *Critical Sepsis* | 20.487 [2.798, 149.987] | **0.0029** | 87.817 [1.511, 5103.024] | **0.0308** |
| **Time point** |  |  |  |  |
| *3 months* | Ref |  | Ref |  |
| *6 months* | 0.817 [0.352, 1.898] | 0.6385 | 0.649 [0.233, 1.802] | 0.4062 |
| *12 months* | 1.201 [0.507, 2.847] | 0.6778 | 0.743 [0.265, 2.085] | 0.5725 |
| **Sex** |  |  |  |  |
| *Male* |  |  | Ref |  |
| *Female* |  |  | 99.795 [2.415, 4123.066] | **0.0153** |
| **Obesity** |  |  |  |  |
| *No* |  |  | Ref |  |
| *Yes* |  |  | 3.754 [0.324, 43.569] | 0.2902 |
| **Smoking status** |  |  |  |  |
| *Never smoked* |  |  | Ref |  |
| *Smoke(d)* |  |  | 0.977 [0.112, 8.564] | 0.9836 |
| **Duration symptoms** |  |  |  |  |
| *Less than 1 week* |  |  | Ref |  |
| *More than 1 week* |  |  | 5.851 [0.466, 73.399] | 0.1710 |
| **Ethnicity** |  |  |  |  |
| *White* |  |  | Ref |  |
| *Non-White/ Unknown* |  |  | 3.170 [0.207, 48.509] | 0.4072 |
| **Vaccination** |  |  |  |  |
| *No* |  |  | Ref |  |
| *Yes* |  |  | 1.056 [0.027, 42.061] | 0.9769 |
| **Wave** |  |  |  |  |
| *1st* |  |  | Ref |  |
| *2nd* |  |  | 7.897 [0.451, 138.428] | 0.1573 |
| *3rd* |  |  | 2.120 [0.037, 122.849] | 0.7167 |
| **ge** |  |  |  |  |
| *<40 yrs* |  |  | Ref |  |
| *40-60 yrs* |  |  | 4.915 [0.156, 155.031] | 0.3659 |
| *>60 yrs* |  |  | 12.594 [0.180, 880.774] | 0.2425 |
| **Clustering Symptoms** |  |  |  |  |
| *Cluster 1* |  |  | Ref |  |
| *Cluster 2* |  |  | 4.613 [0.355, 59.968] | 0.2427 |
| **Clustering Comorbidities** |  |  |  |  |
| *Cluster 1* |  |  | Ref |  |
| *Cluster 2* |  |  | 0.128 [0.011, 1.478] | 0.0996 |

**Table 4.2:** Mood disorder score. Odds ratio estimates and 95% confidence intervals, along with the raw p-value from the corresponding Wald z-test, are presented for both unadjusted (left) and adjusted (right) logistic regression. These analyses aim to predict the presence or absence of the clinical endpoint of interest as a function of severity alone (unadjusted analysis) or as a function of severity plus additional potential confounders (adjusted analysis).

| *Factor* | *Unadjusted analysis* |  | *Adjusted analysis* |  |
| --- | --- | --- | --- | --- |
|  | *Odds reference [95% CI]* | *P-val.* | *Odds reference [95% CI]* | *P-val.* |
| *Ref* | 0.903 [0.361, 2.258] | 0.8273 | 7.647 [0.063, 932.528] | 0.4065 |
|  | *Odds ratio [95% CI]* | *P-val.* | *Odds ratio [95% CI]* | *P-val.* |
| **Severity** |  |  |  |  |
| *Mild/ Moderate/ Severe* | Ref |  | Ref |  |
| *Critical ARDS* | 0.872 [0.195, 3.895] | 0.8573 | 0.449 [0.081, 2.477] | 0.3580 |
| *Critical Sepsis* | 6.903 [1.534, 31.063] | **0.0118** | 2.915 [0.527, 16.116] | 0.2202 |
| **Time point** |  |  |  |  |
| *3 months* | Ref |  | Ref |  |
| *6 months* | 0.854 [0.397, 1.839] | 0.6875 | 0.976 [0.424, 2.245] | 0.9546 |
| *12 months* | 1.373 [0.623, 3.025] | 0.4311 | 1.336 [0.567, 3.149] | 0.5081 |
| **Sex** |  |  |  |  |
| *Male* |  |  | Ref |  |
| *Female* |  |  | 8.300 [1.726, 39.921] | **0.0083** |
| **Obesity** |  |  |  |  |
| *No* |  |  | Ref |  |
| *Yes* |  |  | 3.231 [0.664, 15.718] | 0.1462 |
| **Smoking status** |  |  |  |  |
| *Never smoked* |  |  | Ref |  |
| *Smoke(d)* |  |  | 1.085 [0.277, 4.252] | 0.9067 |
| **Duration symptoms** |  |  |  |  |
| *Less than 1 week* |  |  | Ref |  |
| *More than 1 week* |  |  | 0.888 [0.223, 3.541] | 0.8666 |
| **Ethnicity** |  |  |  |  |
| *White* |  |  | Ref |  |
| *Non-White/ Unknown* |  |  | 1.763 [0.359, 8.668] | 0.4851 |
| **Vaccination** |  |  |  |  |
| *No* |  |  | Ref |  |
| *Yes* |  |  | 1.139 [0.118, 11.014] | 0.9104 |
| **Wave** |  |  |  |  |
| *1st* |  |  | Ref |  |
| *2nd* |  |  | 0.956 [0.176, 5.200] | 0.9589 |
| *3rd* |  |  | 0.206 [0.016, 2.613] | 0.2230 |
| **Age** |  |  |  |  |
| *<40 yrs* |  |  | Ref |  |
| *40-60 yrs* |  |  | 0.307 [0.043, 2.191] | 0.2389 |
| *>60 yrs* |  |  | 0.200 [0.021, 1.915] | 0.1627 |
| **Clustering Symptoms** |  |  |  |  |
| *Cluster 1* |  |  | Ref |  |
| *Cluster 2* |  |  | 1.658 [0.391, 7.025] | 0.4927 |
| **Clustering Comorbidities** |  |  |  |  |
| *Cluster 1* |  |  | Ref |  |
| *Cluster 2* |  |  | 0.231 [0.051, 1.047] | 0.0573 |

In the cohort overall, the OR of being alive at discharge (logistic regression model) was reduced in patients with the most severe disease and this association remained when adjusted for ethnicity, age and comorbidities as well as all other covariates in the table (Table 3.6). Of the other demographic factors recorded, including vaccination status pre-admission there was no significant additive impact over admission COVID-19 severity score on OR of being alive at discharge. In the time to event analysis, the hazard ratio (HR) of death was increased in patients with the most severe disease compared to patients with mild or moderate disease (Table 3.7). In the adjusted analysis, with an additive impact of membership of comorbidities cluster 1, positive vaccination status and shorter time to admission (Table 3.7, less vs. greater than 1 week) were associated with significantly increased HR.

### Clinical Recovery to 12 months post admission

433 follow up visits were made by 231 patients at: 3 months (N=183), 6 months (N=131) and 12 months (N=128) following hospital discharge (or recruitment visit for patients recruited in the community). 68 Patients had 1 visit, 115 had 2 visits and 48 had 3 visits. Admission to Discharge CRFs were matched with Follow up CRFs (see Supplementary Files 1.3-1.4) to evaluate recovery from COVID and how recovery related to the severity of initial COVID presentation and the demographic of the cohort.

Of those patients attending follow up, persisting COVID-19 related symptoms were reported in over 80% of individuals with neurocognitive and neuromuscular problems and breathlessness most reported (Figure 3.1). Over 50% of patients reported at least 3 symptoms that they related to their COVID-19 infection with over 40% of individuals reporting symptoms consistent with a degree of mood disorder. Prevalence of symptoms differed over time since discharge for some but not all parameters. The demographic of the cohorts varied by follow up visit and the number of follow up visits by each patient was variable, potentially confounding interpretation of trajectory of recovery over time. A matched pair analysis for 3-6M visits (N=100) and 3-12M visits (N=90) was performed as is shown in Figure 3.2. A significant improvement in MRC breathlessness, and Rockwood frailty index was seen by 12 months post discharge, however there was no significant change in Mood disorder score and number of COVID-19 symptoms reported.

Women were more likely to report persistent COVID-19 symptoms in 12 month follow up. The odds ratio for many symptoms was greater in women than men and this reached significance for symptoms of myalgia, cognitive problems, loss of taste, gastrointestinal symptoms of constipation, loose stools, abdominal pain and dizziness (Figure 3.3). There was no significant difference in COVID-19 severity score at recruitment by gender in this follow up cohort (p=0.33, data not shown).

To determine the relationship between acute COVID-19 severity score and demographic factors that may predispose patients to persistent symptoms of breathlessness and of mood disorder, we generated the adjusted odds ratios (ORs) for symptoms. We found that female gender and critical sepsis (compared to mild/moderate/severe COVID-19) were significantly associated with breathlessness (Table 3.1). Female gender was also significantly associated with mood disorder (Table 3.2).

## Discussion

This study describes the clinical course of a cohort of patients with predominantly severe COVID-19 (including 89 patients treated with ECMO) through the first 3 waves of the COVID-19 pandemic in England from pre-admission symptoms to recovery to 12 months and identifies the demographic factors associated with severe disease which independently associated with complications of acute disease, survival and persistent COVID-19 symptoms. Our cohort was more severe than in many published studies with nearly 60% requiring intensive care and 21% receiving ECMO. We found that the acute course was often prolonged extending to months in many individuals, and complicated predominantly by thrombotic and renal events. Whilst most inpatients survived to discharge, 22% of the cohort died. COVID-19 disease severity at admission was independently associated with four of the five major complications reported: thrombotic events, neurological events, renal complications and pneumothorax. These associations remained significant after adjusting for pre-admission demographic factors. Renal complications were also associated with obesity and admission in the first wave (compared to second wave) of the pandemic.

Understanding the demographic factors that predict COVID disease severity and acute clinical course is important for health service planning, informed triage at admission, understanding disease mechanisms and developing new therapies. We, as others have reported, found that the distribution of disease severity differs by age, gender, obesity, ethnicity, heart disease, immune suppressant medication, admission reason, wave of admission, vaccine received and vaccination status (Table 1.1) and that disease severity score was a significant risk factor for four of the five major complications reported: thrombotic events, neurological events, renal complications and pneumothorax, and also for survival (Tables 2.1-2.7).

We were interested to explore whether there were preadmission factors that may be helpful in predicting risk of acute complications including survival. Of the demographic factors assessed, we found that obesity was associated with increased risk of acute renal complications. Page-Wilson et al, 2021 also found a slight increased OR of obesity associating with acute renal injury {30}. The reasons for this association are likely to be multifactorial as discussed by Popkin et al in 2020 {31} relating to premorbid renal impairment and regulation of the immune response to SARS-CoV-2. Obesity may associate with a heightened pre-admission pro-inflammatory state as determined by urokinase plasminogen activator receptor (suPAR) concentration which has been reported to be a key mediator between obesity and poor outcome from COVID-19 {32}. Detailed serum cytokine analysis of patients with acute severe COVID has shown distinct inflammatory profiles between obese and non-obese patients segregating with higher prevalence of liver/kidney and lung damage respectively {33}. In addition, there may be a direct association with glomerular damage and the metabolome associated with obesity. It has been shown that SARS-CoV-2 causes a direct glomerular injury, collapsing glomerulopathy (CG) and that GC is strongly associated with apolipoprotein L1(APOL-1) risk variants which themselves are associated with obesity risk {34}.

We also looked at whether the pattern of preadmission symptoms might also be predictive of acute clinical disease course. Identifying real independent associations of patient reported symptoms with acute clinical course of disease is extremely challenging due to the high numbers of potentially confounding variables including the reliability of symptom reporting. The biological mechanisms that may explain the association we report of thrombotic and renal complications with patients who cluster to a narrower breadth and respiratory predominant symptoms likely reflects a bias in dominant symptom reporting in those with most severe disease, noting that many of our patients were recruited in critical care with pre-COVID symptoms collated from medical records. Other studies have reported an association between COVID related GI disturbance and acute clinic course {35, 36, 37}. We did not see this. No other preadmission symptoms as far as we are aware have been found to predict risk of acute disease complications independent of COVID severity score.

Being admitted in the first wave of the pandemic compared to the second wave was also a risk factor for renal complications however there was no difference between first and third wave, despite third wave patients generally having milder disease (Table 1a). Higher risk of renal complication in wave 1 compared with wave 2 may reflect a combination of the learning and changes in medical management of patients with severe COVID-19 as the pandemic evolved. Early reports from China suggested that there was no risk of AKI in patients hospitalised with COVID {38} however subsequent studies from Europe and the USA as summarise by Hilton et al in 2022 reported a greatly increased risk with meta-analyses reporting renal complications in 5-40% of patient hospitalised with COVID {39}. This reduction in risk of acute kidney injury (AKI) over the first 2 waves that we report may reflect the increased awareness of risk of AKI and changes in fluid management policy in subsequent waves that reduced the risk of AKI over time as discussed by Jewell at al., who reported reducing incidence of AKI over time in wave 2 of the UK pandemic in 2 London hospitals {40}. The similar levels of renal complications in wave 3 patients compared with wave 1 is difficult to explain and may be due to inadequate power in the smaller wave 3 cohort to test for wave 3 as an independent confounding variable.

In our study, 6% of our hospitalised cohort developed pneumothorax. This risk was increased in patients with the most severe admission score and more rapid progression from symptom onset to admission. Whilst the prevalence of pneumothorax in patients hospitalised with COVID is reported to be relatively low at 0.3%, it is known to be much greater in patients with severe disease requiring ventilatory support. The association of pneumothorax with shorter time to admission likely reflects a more aggressive disease course to needing ventilatory support, a key admission requirement at the time of the study. Notably, pneumothorax did not associate with comorbidities or smoking status, as has been found in previous systematic reviews reporting that pneumothorax did not associate with pre-morbid lung disease {41}. We found no association with gender or age supporting the assertion by Martinelli et al {42} that the increased prevalence of pneumothorax in males may be attributed to more severe COVID-19 disease.

Neurological complications were reported in 9% of our cohort and whilst the adjusted OR for neurological events increased a little when demographic factors were considered, we found no independent risk association of demographic factors. This may reflect the power of our study to detect effect noting that meta-analysis including larger cohorts have reported ischaemic stroke to have a prevalence of ¬ 2% {43} in hospitalised patients increasing with comorbidities including hypertension, hyperlipidaemia and diabetes and with male gender. We did not differentiate between haemorrhagic and ischaemic stroke noting that both occurred in our cohort. There is little data reported on risk association with haemorrhagic stroke and identifying demographic/pre-admission predictor of risk is complicated by inpatient care, anticoagulant treatment is routinely used to protect against the more common thrombotic complication of pulmonary embolism. COVID is known to be associated with cardiovascular events including arrhythmic, myocarditis and acute thrombosis. The risk is highest in the acute phase. Cardiac complications were reported in 9% of our cohort and did not associate either with COVID-19 severity score or our patient demographic. This contrasts with a careful systematic review of cardiovascular disease in COVID {44}, which reports that COVID disease severity is associated with increased risk of pulmonary embolism, ischaemic stroke and myocardial infarction. In this study, a gender bias in risk was found with pulmonary embolus (PE) being more common in men and myocardial infarction (MI) in women; the impact of age was less clear. Inadequate size effect may explain this discrepancy in results however other confounding variables may be relevant. We did not differentiate patients admitted presenting with an acute cardiac event and those admitted with COVID respiratory disease who subsequently developed cardiac complications and as such our study may not have the granularity to identify cardiovascular disease as a preadmission risk factors with confidence.

### Survival

We used two types of analysis, logistic regression and survival analysis, to evaluate preadmission factors associated with survival to discharge, to examine survival at time of discharge and time to death respectively. In the first analysis, increased COVID severity (critical sepsis), older age (>60 years vs<40 years old), presence of comorbidities and being of unassigned ethnicity impacted was associated with increased probability of death before discharge. The latter highlights the deficit in ethnicity recording in medical records and the challenge of assigning correct ethnicity in the intensive care setting. In the second analysis, increased COVID severity (critical sepsis), a positive vaccination status, shorter duration of symptoms (<1 week) prior to admission, and presence of comorbidities are associated with increased hazard of death. In contrast to the first analysis (logistic regression), which examines probability of death before discharge, this statistical approach (survival analysis) compares risk of death at any point, and accounts for each patient’s length of time in the study. These data corroborate other studies including the large multicentre ISARC-4C study by Docherty et al in 2020 reporting that patients of older age, male gender, with co-morbidities were less likely to survive hospitalisation {45}.

Of our results, the impact of the wave of the pandemic and vaccination status on survival and time to death warrants some discussion. These data emphasise that whilst new variants of COVID have different virulence patterns and vaccination largely protects against severe infection, for patients who require hospitalisation for COVID, mortality remains high. Detailed serological studies are in process to determine whether those individuals vaccinated prior to admission had primary or secondary vaccine failure.

### Recovery

Patients were followed up at regular intervals to 12 months post discharge. Multiple COVID-19 symptoms persisted in many patients and whilst breathlessness improved by 12 months post discharge, symptoms of mood disorder did not. In contrast to acute complications, breathlessness was the only symptom associating with COVID-19 acute disease severity score. Of the demographic factors assessed, female gender was also associated with persistent breathlessness, and this was at a similar OR to, and independent of COVID-19 disease severity on admission. Female gender also associated with persistent symptoms of mood disorder and with neurological and abdominal symptoms. Gender was the only demographic factor associated independently with recovery. These finding corroborate the numerous case series and systematic reviews now published including the large multicentre UK PHOSP study {46} showing that life changing sequelae of COVID are common, complex, unrelated to acute disease severity, predominantly gender biased and may not improve at least over the first year after hospital discharge.

### Strengths and Weaknesses

Our study, although multicentre and prospective, was relatively small and biased to patients with very severe COVID, many requiring prolonged intensive care. The study design adapted to clinical and practical constraints in implementation as the pandemic evolved and it was focused on identifying clinical correlates of disease severity in our cohort to be considered in serological studies exploring the humoral immune correlates of protection. As such this might limit the generalisability of our findings as applied to all hospitalised patients. However as systematic reviews and meta-analysis over the years since the pandemic have shown, results of studies such as ours are generally validated in larger more mixed cohorts from similar geographical areas. The prospective nature of our study, from pre-admission symptoms, acute disease to recovery to 12 months with matched pair analysis provides some confidence in associating clinical course with demographic and pre-admission risk factors; however, there may be more associations that the study was inadequately powered to identify. Similarly, whilst many potentially confounding variables were considered in our analysis, the therapies given during hospital stay were many and diverse such that it was not possible to analyse their contribution to outcome. Mortality following admission with COVID-19 may be under-represented as we did not evaluate mortality post discharge which has recently been reported to be 5% by systemic review and metanalysis of longer-term post-hospital mortality {47}.

In the last 12 months to November 2024 in the UK, an average of 322 patients are hospitalised with COVID each day and 8.5% of deaths have COVID listed as a contributing cause {48}. This cohort of patients and mostly vaccinated (in accordance with national strategy), infected with further evolved variants of the SARS-CoV2 virus, and have comorbidities more likely to be associated with both primary and secondary vaccine failure {49, 50}. As such, the applicability of our findings to patients currently hospitalised with severe COVID is unclear. Ongoing prospective multicentre studies are required to ensure that the extensive learning achieved in the early waves of the pandemic continues to evolve, remaining applicable in real time for this rapidly evolving disease.

## Data Availability

All data produced in the present study are available upon reasonable request to the authors

## Acknowledgements

We thank the RPH Foundation Trust COVID-19 Research and Clinical teams including the Papworth Trials Unit Collaboration (PTUC), and the NIHR Study Support Service LCRN Eastern Core Team for supporting recruitment to this study, HCWs and Outpatients who participated in this study. We give particular thanks to Philip Noyes, Claire Matthews; Allison Doel; Kitty Paques; Jamie Pack; Lucie Garner; Thomas Devine from the PTUC for their invaluable help in supporting all aspects of delivering the study; Janet Piggott, Kim Fell, Sara Thompson and Emma Marsh of the East of England Regional Research Delivery Network in supporting study set up and facilitating multi-site engagement throughout the study; We thank Andrew Barlow and Chiara Ellis at West Hertfordshire NHS Trust; Abigail Crew, Nicola Lancaster, Amy Sanderson at Barnsley Hospital NHS Trust; Carine Cruz, Pearl Baker and Sheena Lim at East and North Herts NHS Trust We acknowledge support from the NIHR/UKRI grant, COV0170 Humoral Immune Correlates from COVID-19 (HICC) based at the Lab of Viral Zoonotics, University of Cambridge.

## Declarations

All authors declare no competing interests.

## Supplementary Files

Supplementary File 1: 1.1-1.4. Clinical Reference Forms

Supplementary Figures: S1.1-1.2:

Figure S1.1 Demographic profile by COVID-19 severity: missing data analysis.

Figure S1.2 Clustering by Symptoms and Clustering be co-morbidities.

