## supplementary file 1.1 for "Defining the pre-admission factors that modify risk of acute complications, survival and long-term recovery from COVID-19"

Study ID [ ][ ][ ] – [ ][ ][ ] – [ ]  DOB [ ][ ] Day [ ][ ][ ] Month [ ][ ] Year  Initials [ ][ ][ ]

### HICC
### Screening

| – Date | Day | Month | Year |
| --- | --- | --- | --- |

#### Study cohort

| | | |
|---|---|---|
| Study cohort | Staff - Acute COVID19 symptoms | ☐ |
| | Staff - Healthy control/history of mild COVID19 | ☐ |
| | COVID19 Patients - recruited as in patient | ☐ |
| | COVID19 Patients - recruited as outpatient | ☐ |
| | Banked samples and anonymised data only (for patients who have not consented) | ☐ |

#### Inclusion criteria *(Not required for cohort: Banked samples and anonymised data only)*

| | | |
|---|---|---|
| Healthy Controls and Patients with history of mild/moderate/severe COVID19 disease | Yes ☐ | No ☐ |
| Informed consent or Consultee Declaration obtained prior to sample collection. | Yes ☐ | No ☐ |

#### Exclusion criteria *(Not required for cohort: Banked samples and anonymised data only)*

| | | |
|---|---|---|
| Unwilling to provide informed consent | Yes ☐ | No ☐ |
| Unwilling to give a blood or appropriate sample | Yes ☐ | No ☐ |

#### Eligible

- Is the patient eligible? Yes ☐ No ☐

*If yes:*

- Date of consent *(Not required for cohort: Banked samples and anonymised data only)* ☐☐ Day ☐☐☐ Month ☐☐ Year

#### Participant details

- Sex: Male ☐ Female ☐
- Ethnicity

| | | | |
|---|---|---|---|
| White British | ☐ | Bangladeshi | ☐ |
| White Irish | ☐ | Other Asian | ☐ |
| Other White | ☐ | Black Caribbean | ☐ |
| Mixed White and Black Caribbean | ☐ | Black African | ☐ |
| Mixed White and Black African | ☐ | Black other | ☐ |
| Mixed White and Asian | ☐ | Chinese | ☐ |
| Other Mixed | ☐ | Not reported | ☐ |
| Indian | ☐ | Other | ☐ |
| Pakistani | ☐ | | |
