## Supplementary file 1.2 for "Defining the pre-admission factors that modify risk of acute complications, survival and long-term recovery from COVID-19"

Study ID [ ][ ][ ] – [ ][ ][ ] – P DOB [ ][ ] Day [ ][ ][ ] Month [ ][ ] Year Initials [ ][ ][ ]

### HICC
### COVID19 Patient Baseline

| | |
|---|---|
| – Date | Day Month Year |

| Risk factors | | | | |
|---|---|---|---|---|
| — Chronic/respiratory diseases (excluding asthma) | Yes | ☐ | No | ☐ |
| — Asthma requiring medication | Yes | ☐ | No | ☐ |
| — Chronic/congenital heart disease | Yes | ☐ | No | ☐ |
| — Hypertension | Yes | ☐ | No | ☐ |
| — Chronic renal disease | Yes | ☐ | No | ☐ |
| — Chronic liver disease | Yes | ☐ | No | ☐ |
| — Chronic neurological disease | Yes | ☐ | No | ☐ |
| — Diabetes requiring insulin, oral hypoglycaemic drugs or diet controlled | Yes | ☐ | No | ☐ |
| *If yes:* | | | | |
| — Specify diabetes type | Type I | ☐ | Type II | ☐ |
| — Immunosuppression (treatment related) | Yes | ☐ | No | ☐ |
| — Immunosuppression (disease related) | Yes | ☐ | No | ☐ |
| — Serious mental illness (including but not restricted to: schizophrenia and psychotic disorders, bipolar disorder, eating disorders, severe depression, personality disorders) | Yes | ☐ | No | ☐ |
| — Dementia | Yes | ☐ | No | ☐ |
| — Obesity (clinically apparent) | Yes | ☐ | No | ☐ |
| *If yes:* | | | | |
| — BMI | 30-39.99 | ☐ | >39 | ☐ |
| — Pregnancy | Yes | ☐ | No | ☐ |
| — Prematurity (<37 weeks GA) | Yes | ☐ | No | ☐ |
| — Travel in 14 days before disease onset | Yes | ☐ | No | ☐ |
| *If yes:* | | | | |
| — Specify destination(s) | | | | |
| — Works as a healthcare worker | Yes | ☐ | No | ☐ |
| — Other risk factor | Yes | ☐ | No | ☐ |
| *If yes:* | | | | |
| — Specify details | | | | |

| Smoking status | | | | | | |
|---|---|---|---|---|---|---|
| — Smoking status | Current smoker | ☐ | Ex-smoker | ☐ | Never smoker | ☐ |
| *If Current or Ex-smoker:* | | | | | | |
| — Number of years smoked | | | | | ☐☐ | years |
| — Average number of cigarettes per day | | | | | ☐☐ | cigarettes |

Study ID [ ][ ][ ] – [ ][ ][ ] – P   DOB [ ][ ] Day [ ][ ][ ] Month [ ][ ] Year   Initials [ ][ ][ ]

#### COVID19 swab and symptoms

– Swab/Specimen date: Day [ ][ ] Month [ ][ ][ ] Year [ ][ ]

– Type of specimen
- Nasal/throat swab [ ]
- Nasopharyngeal/nasal aspirate [ ]
- Sputum [ ]
- Tracheal aspirate [ ]
- Broncho-alveolar lavage [ ]
- Other [ ]
- Unknown [ ]

*If other:*

– Specify specimen type [ ]

– Has the patient had symptoms of COVID19? Yes [ ] No [ ]

*If yes:*

Which of the following symptoms has the patient had:

| | | |
|---|---|---|
| – Anosmia (lost or changed sense of smell) | Yes [ ] | No [ ] |
| – Fatigue | Yes [ ] | No [ ] |
| – Myalgia (pain in a muscle or group of muscles) | Yes [ ] | No [ ] |
| – GI (gastrointestinal infections) | Yes [ ] | No [ ] |
| – Pharyngitis (sore throat) | Yes [ ] | No [ ] |
| – Cough | Yes [ ] | No [ ] |
| – Dyspnoea (difficult or laboured breathing) | Yes [ ] | No [ ] |
| – Fever | Yes [ ] | No [ ] |
| – Headache | Yes [ ] | No [ ] |
| – URTI (upper respiratory tract infection) | Yes [ ] | No [ ] |
| – Other | Yes [ ] | No [ ] |

*If yes:*

– Please specify [ ]

– Date of first symptoms: Day [ ][ ] Month [ ][ ][ ] Year [ ][ ]

*If full date not known, please enter Month-Year*

– Duration of symptoms   1 week or less [ ]   More than 1 week [ ]

#### Severity scoring system

– Severity scoring system

- [ ] Mild
- [ ] Moderate pneumonia
- [ ] Severe pneumonia (RR>30 or O2 requirement)
- [ ] Critical ARDS (PaO2/FiO2<300)
- [ ] Critical Sepsis (includes embolic disease. If patient only has embolic disease score on symptoms and then assign embolic in separate scoring system)
- [ ] Critical Septic shock (vasopressor requirement)

#### Hospitalisation details

– Was the patient admitted to hospital? Yes [ ] No [ ]

*If yes:*

– Date of admission to hospital [ ][ ] Day [ ][ ] Month [ ][ ] Year

– Reason for hospital admission

- [ ] Routine admission
- [ ] Flu
- [ ] RSV
- [ ] COVID-19
- [ ] Acute cardiac event
- [ ] Other

*If other:*

– Specify reason for hospital admission [ ]

– Admitted from

| | | | |
|---|---|---|---|
| Home | [ ] | Private Hospital | [ ] |
| Nursing Home | [ ] | Other UK Hospital | [ ] |
| Residential Home | [ ] | Non UK Hospital | [ ] |
| Temp Accommodation | [ ] | Penal Establishment | [ ] |
| Acute Trust Hospital | [ ] | Unknown | [ ] |
| Mental Health Trust | [ ] | Other | [ ] |

Study ID ☐☐☐ – ☐☐☐ – **P** DOB ☐☐ ☐☐ ☐☐ Initials ☐☐☐

Day | Month | Year

#### Clinical Trials of Investigational Medicinal Products (CTIMPS)

– Is the patient participating in any treatment trials? Yes ☐ No ☐

*If yes:*

– Recovery trial Yes ☐ No ☐

*If yes:*

– Recovery trial arm

- Normal treatment ☐
- Aspirin ☐
- Colchicine ☐
- Baricitinib ☐
- Regeneron ☐
- Dimethyl fumarate ☐
- Other ☐

*If other:*

– Specify Recovery trial arm ☐

– REMAP-CAP Yes ☐ No ☐

*If yes:*

– Specify REMAP-CAP treatment given ☐

– TACTIC-E Yes ☐ No ☐

*If yes:*

– TACTIC-E arm

- EDP1815 ☐
- Ambrisentan and Dapagliflozin ☐
- Standard of care ☐

– TACTIC-R Yes ☐ No ☐

*If yes:*

– TACTIC-R arm

- Baricitinib ☐
- Ravulizumab ☐
- Standard of care ☐

– Is the patient participating in any other CTIMPS? Yes ☐ No ☐

*If yes:*

– Specify other CTIMPS and associated treatments given ☐

#### COVID19 vaccine trials

- Is the patient participating in a COVID19 vaccine trial? Yes ☐ No ☐

  *If yes:*

  - Which COVID19 vaccine trial is the patient participating in? Oxford University ☐ Janssen ☐ Novavax ☐ Other ☐

    *If other:*

    - Please specify ____________

#### COVID19 vaccination

- Has the patient received a vaccination for COVID19? Yes ☐ No ☐

  *If yes:*

  - First dose Yes ☐ No ☐

    *If yes:*

    - First dose date Date ☐☐ Day | ☐☐ Month | ☐☐ Year
    - First dose vaccine Oxford University-AstraZeneca ☐ Moderna ☐ Pfizer-BioNTech ☐ Other ☐

    *If other:*

      - First dose specify ____________

  - Second dose Yes ☐ No ☐

    *If yes:*

    - Second dose date Date ☐☐ Day | ☐☐ Month | ☐☐ Year
    - Second dose vaccine Oxford University-AstraZeneca ☐ Moderna ☐ Pfizer-BioNTech ☐ Other ☐

    *If other:*

      - Second dose specify ____________

  - Booster dose Yes ☐ No ☐

    *If yes:*

    - Booster dose date Date ☐☐ Day | ☐☐ Month | ☐☐ Year
    - Booster dose vaccine Oxford University-AstraZeneca ☐ Moderna ☐ Pfizer-BioNTech ☐ Other ☐

    *If other:*

      - Booster dose specify ____________
