## Supplementary file 1.3 for "Defining the pre-admission factors that modify risk of acute complications, survival and long-term recovery from COVID-19"

Study ID | | | | – | | | | – | P | DOB | | | | | | | | | Initials | | |

Day | Month | Year

### HICC
### COVID19 Patient Discharge

– Date

Day | Month | Year

#### Hospitalisation details

— Was the patient admitted to hospital? Yes ☐ No ☐

*If yes, complete the rest of the form*

*If no, end of form.*

#### ICU

— Was the patient admitted to ICU? Yes ☐ No ☐

*If yes:*

— Days in ICU ☐☐☐

#### Respiratory support

— Did the patient require respiratory support? Yes ☐ No ☐

*If yes:*

— Oxygen via cannula or mask Yes ☐ No ☐

— High flow nasal oxygen Yes ☐ No ☐

— Non-invasive ventilation Yes ☐ No ☐

*If yes:*

— Number of days ☐☐☐

— Invasive mechanical ventilation Yes ☐ No ☐

*If yes:*

— Number of days ☐☐☐

— ECMO Yes ☐ No ☐

*If yes:*

— Number of days ☐☐☐

#### Other support

— Did the patient require haemofiltration? Yes ☐ No ☐

— Did the patient require plasmapheresis? Yes ☐ No ☐

— Did the patient require cytosorp? Yes ☐ No ☐

Study ID [ ][ ][ ] – [ ][ ][ ] – P DOB [ ][ ] Day [ ][ ][ ] Month [ ][ ] Year Initials [ ][ ][ ]

#### Complications

- Thrombotic events Yes [ ] No [ ]
- Cardiac events Yes [ ] No [ ]
- Neurological events Yes [ ] No [ ]
- Renal complications Yes [ ] No [ ]
- Pneumothorax Yes [ ] No [ ]
- Secondary bacterial pneumonia Yes [ ] No [ ]

*If yes:*

- Specify secondary bacterial pneumonia

| | |
|---|---|
| Aspergillus [ ] | Mycoplasma [ ] |
| Candida [ ] | Pseudomonas [ ] |
| E-coli [ ] | Staphylococcus aureus [ ] |
| Fungal pneumonia [ ] | Staphylococcus epidermidis [ ] |
| Haemophilus [ ] | Streptococcus [ ] |
| Klebsiella [ ] | Other [ ] |

*If other:*

- Specify other organism [ ]

- Other complications Yes [ ] No [ ]

*If yes:*

- Specify other complications [ ]

#### Treatment

- Treated with systemic corticosteroids? Yes [ ] No [ ]

#### Final outcome

| | |
|---|---|
| — Final outcome | Discharged ☐ |
| | Transferred ☐ |
| | Death ☐ |
| — Date | Day ☐☐ Month ☐☐☐ Year ☐☐ |

*If transferred:*

| | |
|---|---|
| — Transfer destination | Acute Trust Hospital ☐ |
| | Private hospital ☐ |
| | Unknown ☐ |
| | Other ☐ |
| | Other NHS hospital ☐ |
| | Care home ☐ |
| | General ward ☐ |
| | Different ICU/HDU ☐ |

*If death:*

| | |
|---|---|
| — Cause of death | COVID-19 main cause ☐ |
| | COVID-19 underlying cause ☐ |
| | Not COVID-19 related ☐ |
| | Influenza main cause ☐ |
| | Influenza underlying cause ☐ |
| | RSV main cause ☐ |
| | RSV underlying cause ☐ |
| | Other ☐ |
| | Unknown ☐ |
