## Supplementary file 1.4 for "Defining the pre-admission factors that modify risk of acute complications, survival and long-term recovery from COVID-19"

Study ID ☐☐☐ – ☐☐☐ – P DOB ☐☐ ☐☐☐ ☐☐ Initials ☐☐☐

Day | Month | Year

### HICC
### COVID19 Patient Follow-up

| | |
|---|---|
| – Date | ☐☐ / ☐☐☐ / ☐☐ (Day / Month / Year) |
| – Follow-up | 3 month ☐ |
| | 6 month ☐ |
| | 12 month ☐ |
| | Unscheduled visit ☐ |

#### Symptoms on follow-up

*On a scale of 0-5 (where 0 = I do not have this problem and 5 = the symptom is very significant), please rate the following symptoms. Also grade the severity at maximum and in general whether staying the same, getting better or getting worse.*

| | | | | | | |
|---|---|---|---|---|---|---|
| — Breathlessness | 0 ☐ | 1 ☐ | 2 ☐ | 3 ☐ | 4 ☐ | 5 ☐ |
| — Breathlessness max | 0 ☐ | 1 ☐ | 2 ☐ | 3 ☐ | 4 ☐ | 5 ☐ |
| — Breathlessness trajectory | | Same ☐ | | Better ☐ | | Worse ☐ |
| — Cough | 0 ☐ | 1 ☐ | 2 ☐ | 3 ☐ | 4 ☐ | 5 ☐ |
| — Cough max | 0 ☐ | 1 ☐ | 2 ☐ | 3 ☐ | 4 ☐ | 5 ☐ |
| — Cough trajectory | | Same ☐ | | Better ☐ | | Worse ☐ |
| — Fatigue | 0 ☐ | 1 ☐ | 2 ☐ | 3 ☐ | 4 ☐ | 5 ☐ |
| — Fatigue max | 0 ☐ | 1 ☐ | 2 ☐ | 3 ☐ | 4 ☐ | 5 ☐ |
| — Fatigue trajectory | | Same ☐ | | Better ☐ | | Worse ☐ |
| — Sleep | 0 ☐ | 1 ☐ | 2 ☐ | 3 ☐ | 4 ☐ | 5 ☐ |
| — Sleep max | 0 ☐ | 1 ☐ | 2 ☐ | 3 ☐ | 4 ☐ | 5 ☐ |
| — Sleep trajectory | | Same ☐ | | Better ☐ | | Worse ☐ |

#### MRC Breathlessness score

— MRC Breathlessness score

- 1 Not troubled by breathless except on strenuous exercise ☐
- 2 Short of breath when hurrying on a level or when walking up a slight hill ☐
- 3 Walks slower than most people on the level, stops after a mile or so, or stops after 15 minutes walking at own ☐
- 4 Stops for breath after walking 100 yards, or after a few minutes on level ground ☐
- 5 Too breathless to leave the house, or breathless when dressing/undressing ☐

Study ID [ | | ] — [ | | ] — P   DOB [ | ] Day [ | | ] Month [ | ] Year   Initials [ | | ]

| Neurological symptoms | | | | |
|---|---|---|---|---|
| — Headaches | Yes | ☐ | No | ☐ |
| — Sensory disturbance | Yes | ☐ | No | ☐ |
| — Cognition or memory | Yes | ☐ | No | ☐ |
| — Visual disturbance | Yes | ☐ | No | ☐ |
| — Motor weakness | Yes | ☐ | No | ☐ |
| — Focal weakness or numbness | Yes | ☐ | No | ☐ |

| Cardiac symptoms | | | | |
|---|---|---|---|---|
| — Dizziness | Yes | ☐ | No | ☐ |
| — Palpitations | Yes | ☐ | No | ☐ |
| — Chest pains | Yes | ☐ | No | ☐ |

| GI symptoms | | | | |
|---|---|---|---|---|
| — Loose bowels | Yes | ☐ | No | ☐ |
| — Abdominal pains | Yes | ☐ | No | ☐ |
| — Constipation | Yes | ☐ | No | ☐ |

| COVID specific symptoms | | | | |
|---|---|---|---|---|
| — Myalgia | Yes | ☐ | No | ☐ |
| — Anosmia | Yes | ☐ | No | ☐ |
| — Loss of taste | Yes | ☐ | No | ☐ |

| Symptoms suggestive of PE | | | | |
|---|---|---|---|---|
| — Haemoptysis | Yes | ☐ | No | ☐ |
| — Ongoing or worsening SOB | Yes | ☐ | No | ☐ |
| — Leg swelling | Yes | ☐ | No | ☐ |

| Other symptoms | | | | |
|---|---|---|---|---|
| — Skin rash | Yes | ☐ | No | ☐ |
| — Other symptoms | Yes | ☐ | No | ☐ |

*If yes:*

— Specify other symptoms [ ]

#### Return to work

– Has the patient returned to work/would they feel able to return to work if permitted?

- [ ] Full time
- [ ] Part time
- [ ] Not at all
- [ ] Not applicable

#### Psychological wellbeing

– Little interest or pleasure in doing things

- [ ] Not at all
- [ ] Several days
- [ ] More than half the days
- [ ] Nearly every day

– Feeling down, depressed, or hopeless

- [ ] Not at all
- [ ] Several days
- [ ] More than half the days
- [ ] Nearly every day

– Feeling nervous, anxious or on edge

- [ ] Not at all
- [ ] Several days
- [ ] More than half the days
- [ ] Nearly every day

– Not being able to stop or control worrying

- [ ] Not at all
- [ ] Several days
- [ ] More than half the days
- [ ] Nearly every day

Study ID ☐☐☐ – ☐☐☐ – P    DOB ☐☐ (Day) ☐☐☐ (Month) ☐☐ (Year)    Initials ☐☐☐

#### Rockwood (Clinical Frailty Score)

– Rockwood (Clinical Frailty Score)

| Option | |
|---|---|
| 0 - Patient not assessed | ☐ |
| 1 - Very fit | ☐ |
| 2 - Well | ☐ |
| 3 - Managing well | ☐ |
| 4 - Vulnerable | ☐ |
| 5 - Mildly frail | ☐ |
| 6 - Moderately frail | ☐ |
| 7 - Severely frail | ☐ |
| 8 - Very severely frail | ☐ |
| 9 - Terminally ill | ☐ |

#### MOCA

– Montreal Cognitive Assessment completed? Yes ☐ No ☐

*If yes:*

– Montreal Cognitive Assessment score ☐☐

#### 6 minute walk

– 6 minute walk completed? Yes ☐ No ☐

*If yes:*

| Item | Value | Unit |
|---|---|---|
| – 6 minute walk distance | ☐☐☐ | m |
| – BORG Pre | ☐ . ☐ | |
| – BORG Post | ☐ . ☐ | |
| – SpO2 Baseline | ☐☐☐ | % |
| – SpO2 Min | ☐☐☐ | % |
| – HR Rest | ☐☐☐ | BPM |
| – HR Max | ☐☐☐ | BPM |

#### X-Ray

– Was an X-Ray completed? Yes ☐ No ☐

*If yes:*

– Were there any abnormalities? Yes ☐ No ☐

*If yes:*

– Specify abnormalities

#### Routine bloods

– Were routine blood tests done? Yes ☐ No ☐

*If yes:*

Were there any abnormalities in…

| | Yes | No | Not done |
|---|---|---|---|
| – Lymphocyte count | ☐ | ☐ | ☐ |
| – Urea/creatinine | ☐ | ☐ | ☐ |
| – ANA | ☐ | ☐ | ☐ |
| – ANCA | ☐ | ☐ | ☐ |
| – IFN | ☐ | ☐ | ☐ |
| – Immunoglobulins | ☐ | ☐ | ☐ |
| – D-Dimers | ☐ | ☐ | ☐ |
| – Hb1AC | ☐ | ☐ | ☐ |
| – LDH | ☐ | ☐ | ☐ |
| – TAG | ☐ | ☐ | ☐ |
| – CRP | ☐ | ☐ | ☐ |
| – proBNP | ☐ | ☐ | ☐ |

Study ID [ | | ] – [ | | ] – [P] DOB [ | ] [ | | ] [ | ] Initials [ | | ]
Day Month Year

#### COVID19 vaccine trials

– Is the patient participating in a COVID19 vaccine trial? Yes ☐ No ☐

*If yes:*

– Which COVID19 vaccine trial is the patient participating in? Oxford University ☐ Novavax ☐ Janssen ☐ Other ☐

*If other:*

– Please specify [ ]

#### COVID19 vaccination

– Since their last follow-up, has the patient received a vaccination for COVID19? Yes ☐ No ☐

*If yes:*

– First dose Yes ☐ No ☐

*If yes:*

– First dose date Date [ | ] [ | | ] [ | ]
Day Month Year

– First dose vaccine Oxford University-AstraZeneca ☐ Moderna ☐ Pfizer-BioNTech ☐ Other ☐

*If other:*

– First dose specify [ ]

– Second dose Yes ☐ No ☐

*If yes:*

– Second dose date Date [ | ] [ | | ] [ | ]
Day Month Year

– Second dose vaccine Oxford University-AstraZeneca ☐ Moderna ☐ Pfizer-BioNTech ☐ Other ☐

*If other:*

– Second dose specify [ ]

– Booster dose Yes ☐ No ☐

*If yes:*

– Booster dose date Date [ | ] [ | | ] [ | ]
Day Month Year

– Booster dose vaccine Oxford University-AstraZeneca ☐ Moderna ☐ Pfizer-BioNTech ☐ Other ☐

*If other:*

– Booster dose specify [ ]

This page has been intentionally left blank.
