## Supplementary Figure 1.1_1.2 for "Defining the pre-admission factors that modify risk of acute complications, survival and long-term recovery from COVID-19"

**Supplementary Figure S1.1 Demographic profile by COVID-19 severity.** Heatmap displaying the missingness status (red for missing data and grey for available data) for each demographic variable (y-axis) and patient (x-axis). The net sample size and proportion of non-missing data are indicated for each variable.

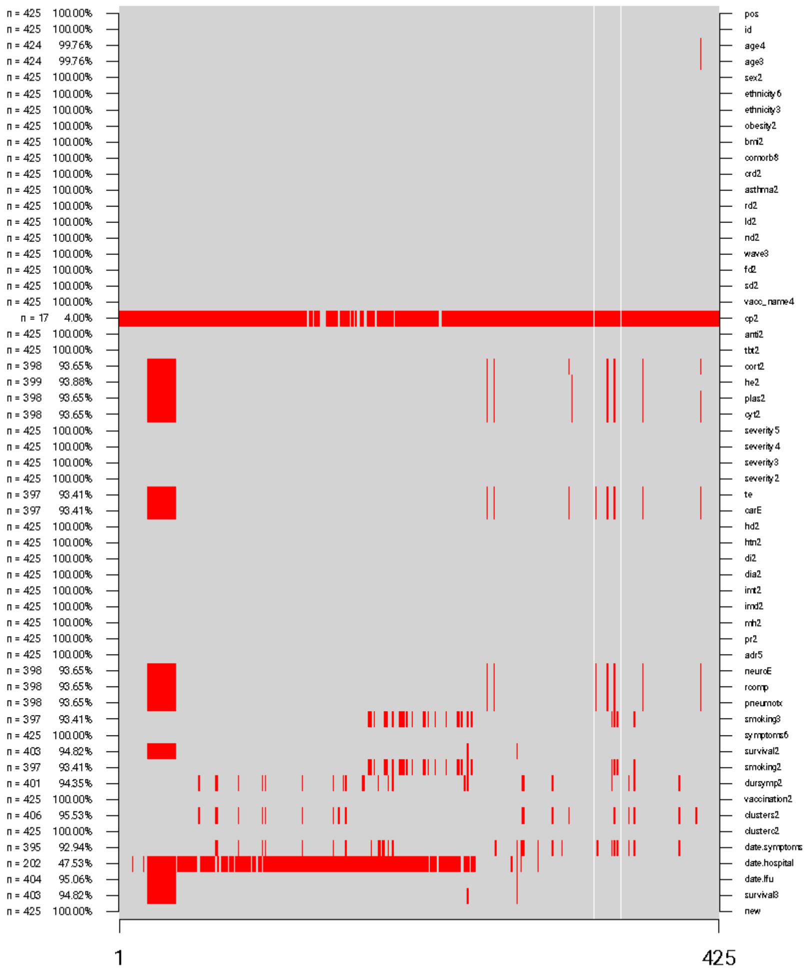

**Supplementary Figure S1.2 Clustering by Symptoms and Clustering be co-morbidities.** Comparison of the probability of observing each symptom (left plot) and comorbidity (right plot) per cluster. For each symptom and comorbidity, stars indicate the significance level of a test for equality of proportions.

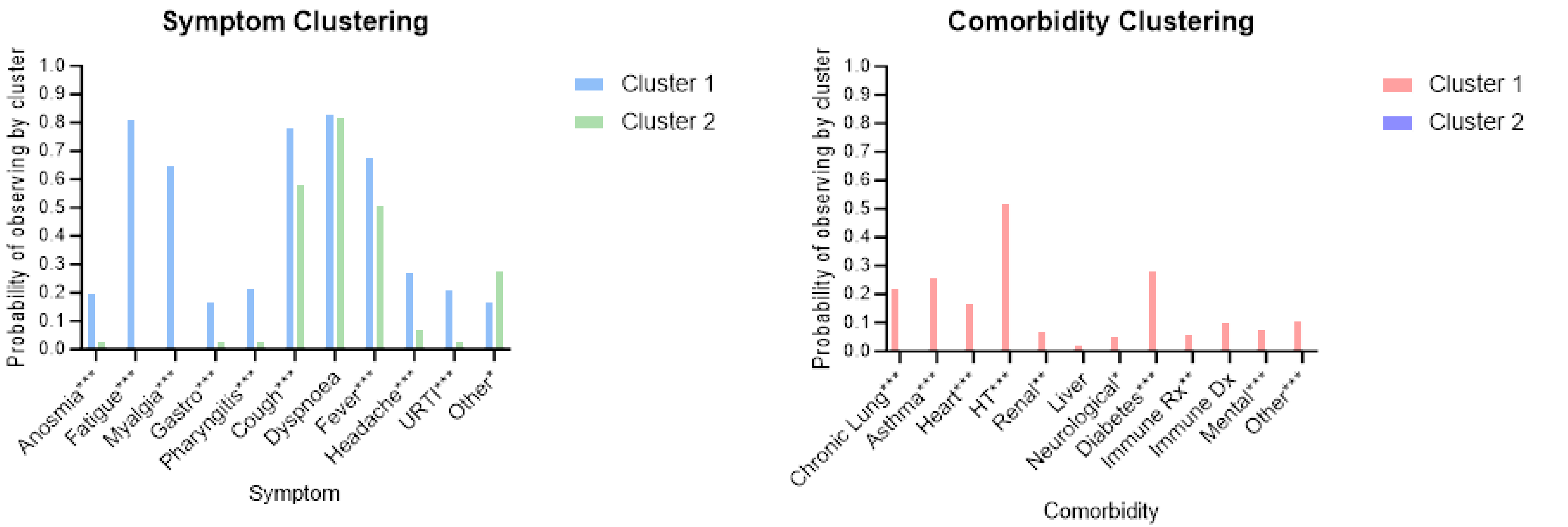
